# Germline haploinsufficiency for the base excision repair gene *MUTYH* causes mutational signature SBS18 in multiple tumour types, specifically leading to an increased risk of colorectal cancer

**DOI:** 10.1101/2025.11.20.25340655

**Authors:** Kitty Sherwood, Juan Fernandez-Tajes, Güler Gül, Steve Thorn, Joseph C. Ward, James F. Wilson, Claire Palles, D. Timothy Bishop, Richard S. Houlston, Malcolm G. Dunlop, Ian P.M. Tomlinson

**Affiliations:** Department of Oncology, Old Road Campus Research Building, University of Oxford, UK; Scotland Cancer Centre, Institute of Genetics and Cancer, University of Edinburgh, Edinburgh, UK; MRC Human Genetics Unit, University of Edinburgh, Edinburgh, UK; Centre for Genomic and Experimental Medicine, Institute of Genetics and Cancer, University of Edinburgh, UK; Centre for Global Health Research, Usher Institute, University of Edinburgh, Edinburgh, UK; Institute of Cancer and Genomic Sciences, College of Medical and Dental Science, University of Birmingham, Birmingham, UK; Leeds Institute of Medical Research, University of Leeds, Leeds, UK; Division of Genetics and Epidemiology, The Institute of Cancer Research, Sutton, UK

**Keywords:** mutational signatures, hypermutation, base excision repair, oxidative DNA damage, *MUTYH*, cancer risk

## Abstract

The MUTYH base excision repair protein corrects oxidative DNA damage. Bi-allelic germline *MUTYH* mutations cause a rare, Mendelian recessive syndrome of colorectal adenomas, duodenal polyps and colorectal cancer (CRC), in which tumours have excess somatic C:G>A:T mutations and the mutational signature SBS36. Signature SBS18, which resembles SBS36, is common in sporadic CRCs and other cancers. Increased risks of CRC and other cancers have been reported in germline *MUTYH* heterozygotes (mono-allelic mutation carriers, frequency 2-3%), but the existence of these associations and underlying mechanism, have remained controversial. Compared with *MUTYH-*wildtype individuals, CRCs from *MUTYH* heterozygotes had ∼2.5-fold excess of signature SBS18, increased C:G>A:T transversions (including in driver genes) and raised mutation burden. These observations resulted from *MUTYH* haploinsufficiency, rather than somatic loss of the wildtype allele, contrary to previous suggestions. Hypermutation probably begins before cancer initiation. In case-control analysis, we found approximately 1.2-fold elevated risk of CRC in *MUTYH* heterozygotes, causally mediated through increased SBS18. The association between *MUTYH* heterozygosity and SBS18 was also present in many extra-colonic cancers, including other gastrointestinal tumours, but the raised SBS18 did not detectably increase the risk of these cancers. Germline heterozygotes for another base excision repair gene, *MBD4*, showed equivalent associations for SBS1, C:G>T:A mutations and CRC risk. Mutational signatures in cancers can result in part from non-rare germline variation in DNA repair. The specific effect of *MUTYH* heterozygosity on CRC risk plausibly reflects the high baseline levels of oxidative damage and SBS18 activity in the colorectum.

## Introduction

Mendelian cancer syndromes frequently show recessive inheritance, implying that a single functional copy of the gene can provide enough residual function to prevent tumorigenesis. Most recessive cancer syndromes take the form of DNA repair deficiency and/or a tendency to hypermutation [1]. The underlying defects can include a failure to repair chromosomal-scale lesions (e.g. Fanconi anaemia), environmentally-induced lesions (e.g. xeroderma pigmentosum), DNA replication errors (e.g. constitutional mismatch repair deficiency) and ‘spontaneous’ mutations (e.g. deamination of methylcytosine). An ongoing question that applies to all of these syndromes is whether mono-allelic mutation carriers (simple heterozygotes), who are evidently much more common than bi-allelic mutation carriers (homozygotes or compound heterozygotes), are also at increased cancer risk. This is clearly the case for genes such as *BRCA2* [2], the Lynch syndrome loci [3] and *ATM* [4], for which heterozygotes are prone to specific tumour types that overlap partially with those present in the recessive syndrome [5].

Although a haploinsufficiency mechanism is theoretically possible, the increased risk of cancer in carriers of these mono-allelic mutations generally occurs through spontaneous somatic ‘second hits’, in classical tumour suppressor gene fashion [6].

Given that somatic second hits are a proven mechanism of increased cancer risk, it is reasonable to ask whether all mono-allelic carriers of mutations associated with recessive DNA repair deficiency syndrome alleles have a raised risk of cancer, since spontaneous somatic inactivation of the germline wildtype allele can occur through spontaneous loss of heterozygosity or another mechanism. However, strong statistical support for an increased cancer risk in heterozygotes is often hard to obtain: risk allele frequencies can be as low as 0.1% or less; family-based studies are prone to ascertainment bias; and apparent germline heterozygotes may actually carry cryptic second germline mutations. Irrespective of the question of raised cancer risk in DNA repair deficiency heterozygotes *via* the two-hit model, even chance second hits at these loci could, in principle, create null DNA repair genotypes and hence specific, important vulnerabilities. Therapeutic strategies against other DNA repair components, or to promote DNA damage of a specific type, could thus provide a cancer cell-specific reduction in viability. However, supporting evidence for ‘second hits’ in tumours is often lacking, because tumour samples are either unavailable or not analysed fully, or a limited genomic assessment has been performed.

The *MUTYH* (mutY homologue) gene maps to chromosome 1p34.1 and encodes a protein with an important role in the base excision repair (BER) of oxidative DNA damage, removing adenine paired with 8-oxo-guanine, and thus preventing C:G>A:T base substitutions [7]. Approximately 0.05% of the population have bi-allelic, pathogenic germline *MUTYH* variants that cause a constitutional deficiency in the repair of oxidative damage [8]. In European populations, the majority of such individuals are homozygous or compound heterozygous for two missense *MUTYH* variants, p.Gly396Asp and p.Tyr179Cys (ENST00000710952.2), which have individual allele frequencies of about 0.1-1.0% and encode a protein deficient in correcting 8-oxoG:A replication mispairs [9].

Carriers of bi-allelic germline *MUTYH* mutations are predisposed to a high-penetrance recessive syndrome, MUTYH-associated polyposis (MAP), that comprises multiple colorectal adenomas and colorectal carcinomas (CRCs), with a smaller risk of duodenal adenomas and carcinomas. The tumours display mutational signature SBS36 [10–12], which is typified by TCA>TAA and TCA>TAT substitutions, and sometimes the similar signature SBS18, which is found in many sporadic CRCs and is thought to arise from oxidative DNA damage in general (**Supplementary Figure 1**). Tumorigenesis in MAP largely occurs along conventional genetic pathways, *via* an increased burden of somatic C:G>A:T changes in driver genes such as *APC, KRAS* and *SMAD4* [13]. An increased risk of other cancer types in MAP remains unproven.

There is a longstanding, unresolved controversy as to whether the 2-3% of the population who are germline *MUTYH* heterozygotes (mono-allelic mutation carriers) have an increased risk of CRC [14–17]. Studies addressing this question have differed in their sizes, genotyping methods, patient inclusion criteria and their marching of cases and controls, problems may have arisen (*e.g.* owing to the accidental inclusion in studies of undetected bi-allelic mutation carriers). In one such recent study, Barreiro *et al* [18] took an All-cancer approach, in which genetic epidemiological data from TCGA cases [19] and gnomAD controls [20] were combined with analysis of somatic ‘second hits’ at *MUTYH* and mutational signatures in tumours. The analysis concluded that there was an overall excess of cancer in *MUTYH* heterozygotes, driven principally by patients with adrenocortical carcinoma, oesophageal carcinoma, sarcoma, prostate adenocarcinoma and renal clear cell carcinoma. Mechanistically, somatic loss of the wildtype *MUTYH* allele was postulated to lead to loss of MUTYH activity and thus to hypermutation, raised SBS18 or SBS36 activity, and accelerated tumorigenesis. However, of note, *MUTYH* heterozygotes were not predisposed to CRC in that study.

Here, we analyse the features of germline *MUTYH* heterozygotes in the UK 100,000 Genomes Project (100kGP), including their cancer risk and the molecular characteristics of their cancers. We then address similar factors for another BER gene *MBD4.* Our results help to elucidate the magnitude and mechanism of cancer risk in BER gene germline heterozygotes, and indicate that COSMIC mutational signatures can originate from non-rare germline variation in DNA repair.

## Results

### Germline and somatic MUTYH genotypes in 100kGP CRCs

We examined the tumour genomes of 2,513 CRCs that we analysed as part of the 100kGP (**Figure 1**) [21]. We classified patients into five groups, defined by their germline and somatic *MUTYH* status (**Figures 1-3**, **Supplementary Tables 1 & 2, Supplementary Figure 2**):

**Figure 1.**
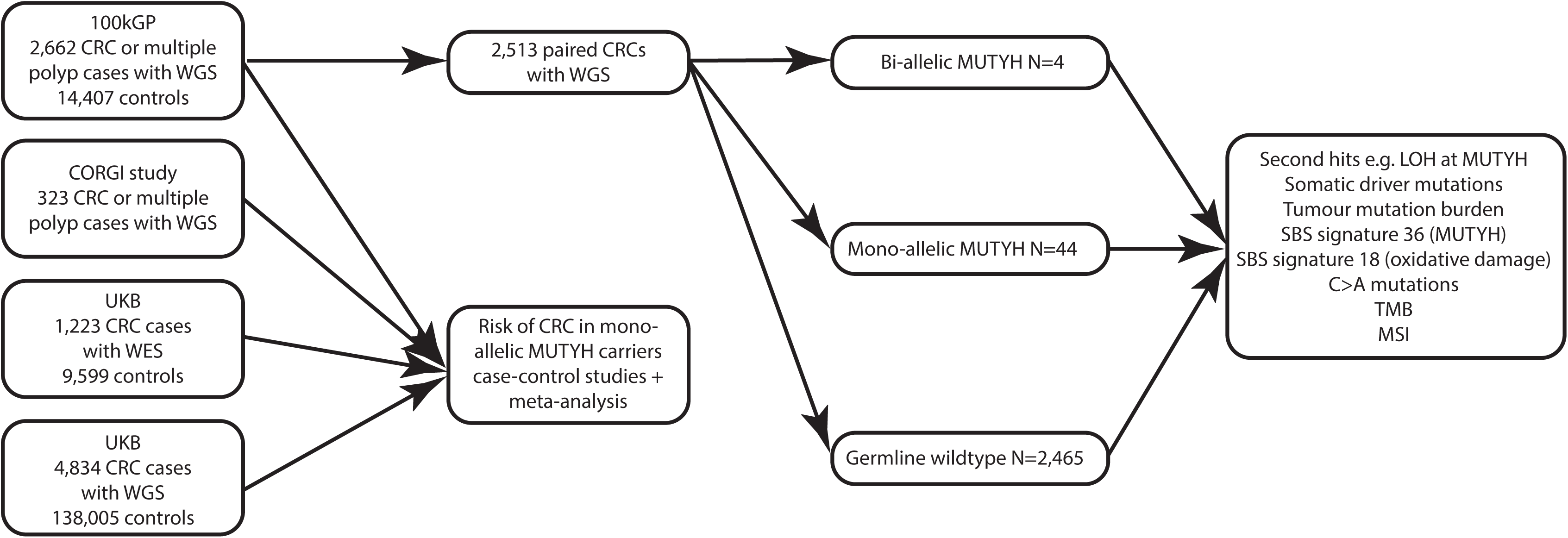
Study design: consort-style diagram of CRC patients included in germline and somatic genetic analyses.

**Figure 2.**
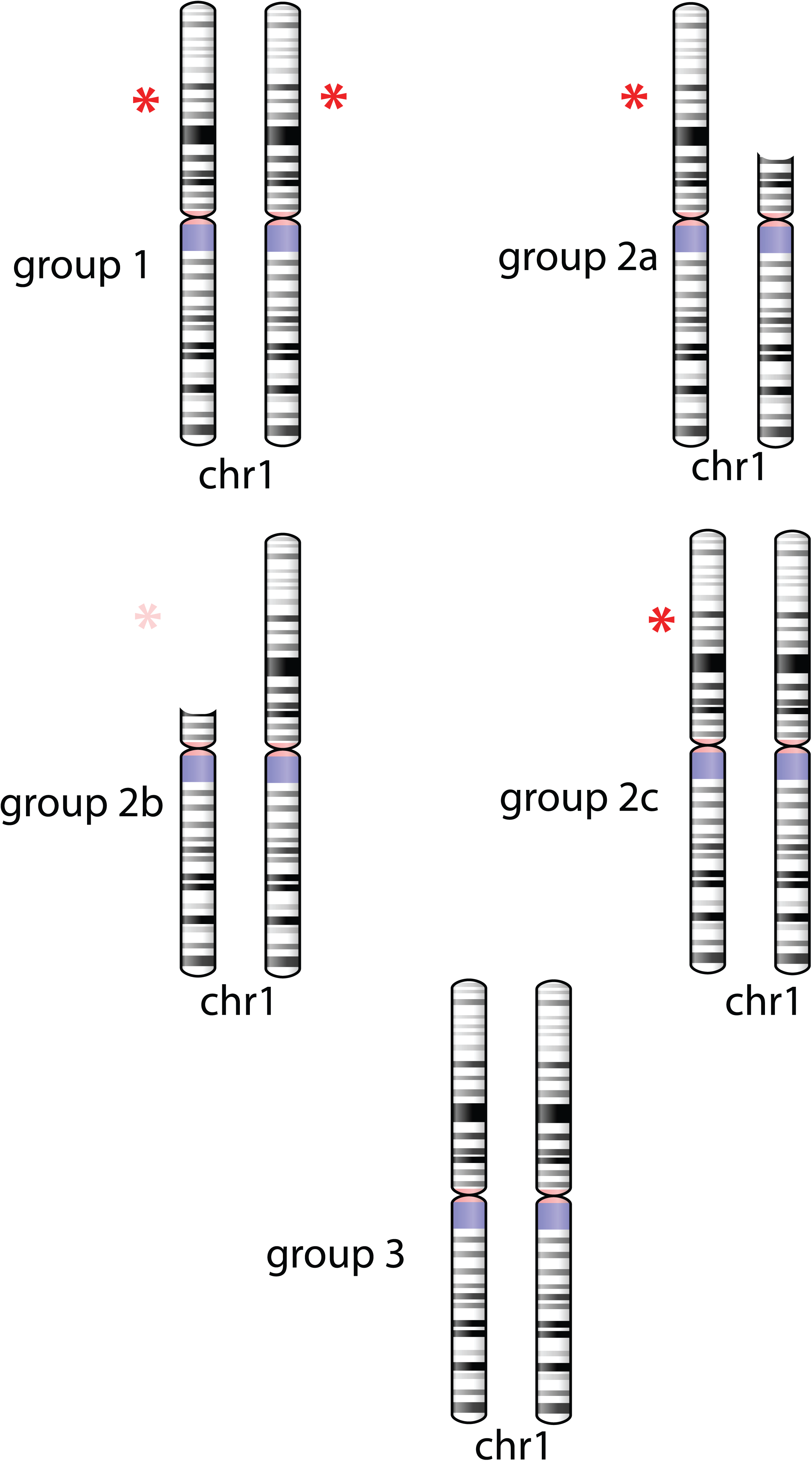
Chromosome 1 ideograms showing the group 1, 2a, 2b, 2c and 3 genotypes in cancers. Germline *MUTYH* mutations are shown by red stars, and somatic second hits by LOH are shown as removing part of the chromosome short arm (copy-loss LOH), although in reality they may instead simply render part of the chromosome homozygous (copy-neutral LOH) in some tumours.

**Table 1.**
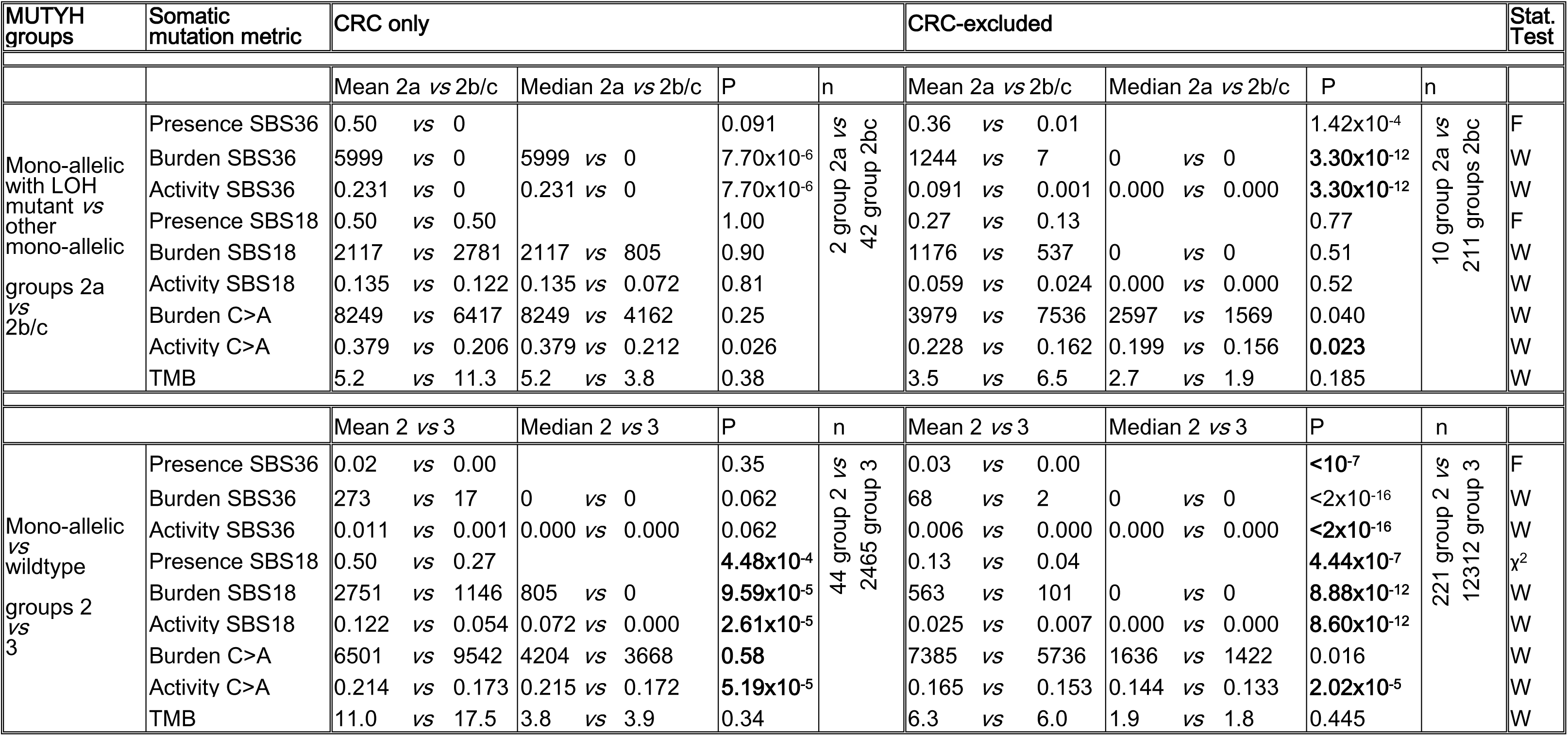
Synopsis of associations between germline MUTYH genotypes and somatic mutation metrics in 100kGP cancers. For signature presence/absence (binary), mean shows the proportion of positive individuals, whilst median has no utility and is not shown. P values are uncorrected and derived from univariable tests (F, Fisher’s exact; χ^2^; W, Wilcoxon). **Bold**, associations remaining significant in multivariable regression. More detailed data are in **Supplementary Tables 1-7 & 13-16**. Note that for CRC in particular, the means for some measures of mutation are higher in group 3 than group 2, whereas the medians are higher in group 2 than group 3, principally due to some MMRd tumours with very high TMB (**Supplementary Tables 1 & 2**).

(1) bi-allelic germline mutations (n=4);

(2a) mono-allelic germline mutation with somatic LOH of the wildtype allele (n=2);

(2b) mono-allelic germline mutation with somatic LOH of the mutant allele (n=5);

(2c) mono-allelic germline mutation with no somatic LOH (n=37); and

(3) wildtype germline (n=2,465).

Somatic mutational processes in tumours were measured in three related ways: (1) *Prevalence* denoted the presence or absence of a mutational signature (e.g. prevalence of SBS36 measured the proportion of cancers with any SBS36 activity); (2) *Burden* measured the absolute numbers of a mutation type in a cancer, such as a specific single base change or mutations assigned to a particular signature; and (3) *Proportional activity* assessed the proportion of all mutations of a specific type (e.g. SBS36 activity was the percentage of mutations assigned to SBS36, relative to all mutations assigned to an SBS signature in that cancer). Prevalence was used to show whether a specific signature was active within a tumour, burden was particularly useful for identifying hypermutant tumours, and activity corrected for background burdens to identify the most active processes in a single tumour. Further details are provided in **Methods**. No tumour had a germline *MUTYH* mutation accompanied by a somatic mutation other than LOH, and there was no evidence of somatic *MUTYH* hypermethylation in a subset of 328 CRCs with EPIC v2 850K CpG methylation data (details not shown).

Groups 2a, 2b and the much larger 2c had similar C:G>A:T, SBS36 and SBS18 metrics (*P>*0.15, Kruskal-Wallis test; **Table 1**, **Supplementary Tables 1** & **2**) and were therefore combined into a single group 2 for subsequent analyses, with the exception of specific investigations of the effects of somatic loss of the germline wildtype *MUTYH* allele (group 2a).

Very few group 3 CRCs had acquired a somatic mutation specifically involving *MUTYH* and none had acquired bi-allelic somatic *MUTYH* mutations. It was common (frequency ∼30%) for MSS CRCs to show heterozygous somatic allelic imbalance encompassing the *MUTYH* locus, but this generally reflected a 2:1 ratio of the parental copies of chromosome 1 in cancers with a near-triploid karyotype, rather than any targeting or loss of *MUTYH.* Specifically, the 4.1% of MSS cancers with either a somatic deletion of 1p or a single, heterozygous somatic mutation had almost the same SBS18 prevalence as cancers with no change of any type at *MUYTH* (OR=1.06, 95%CI 0.780-1.44, *P=*0.765). All these CRCs were therefore analysed as part of the ‘*MUTYH* wildtype’ group 3.

### Associations between germline MUTYH genotype and somatic molecular features of CRCs from the 100kGP

Owing to the reported link between bi-allelic *MUTYH* mutations and SBS36, we searched initially for associations between germline *MUTYH* genotypes and mutational signatures SBS36 and SBS18 in the sporadic CRCs. We also tested for associations with measures of mutational load, specifically C:G>A:T substitution burden and activity, and total mutation burden (using somatic coding variants per megabase (SCVPM) as a measure). For clarity, we summarise the main results here, in **Table 1** and in **Figures 3 and 4**. Full supporting data are provided in **Supplementary Tables 1-8**.

First, compared with all other CRCs, bi-allelic germline *MUTYH* mutations (group 1) were positively associated with all three SBS36 metrics (presence, burden and activity), higher C:G>A:T burden and activity, and higher overall TMB (**Figure 3**). There were no significant associations with SBS18. These results were largely confirmatory of previous studies [8, 22].

**Figure 3.**
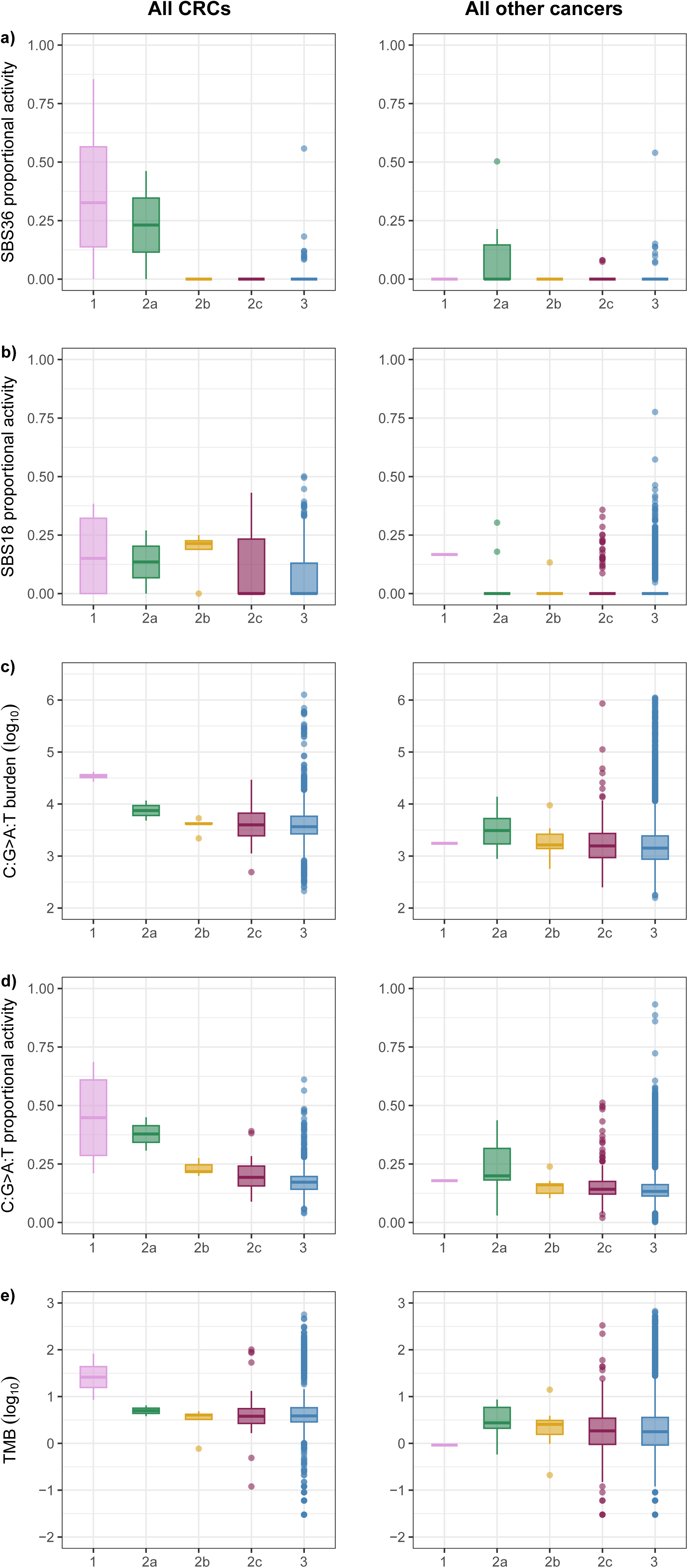
Selected somatic molecular features of tumours in the 100kGP CRC analysis in relation to germline MUTYH status. Group 1, bi-allelic germline mutations; 2a, heterozygous with somatic loss of germline wildtype; 2b, heterozygous with somatic loss of germline mutant; 2c, heterozygous with no somatic mutation; and 3, germline wildtype. (a) SBS36 proportional activity, (b) SBS18 proportional activity, (c) C:G>A:T burden, (d) C:G>A:T proportional activity and (e) tumour mutational burden (measured as SCVPM) in germline MUTYH genotype in 100kGP CRCs (left) and All Other Cancers (right). Log scales are used to aid display in some cases. Significant pairwise univariable associations at P<0.05 in Wilcoxon tests were, in each panel: (a) group 1 *vs* groups 2&3, (b) group 2 *vs* group 3, (c) group 1 *vs* groups 2&3 and (d) both group 1 *vs* groups 2&3, and group 2 *vs* group 3. Details of these associations from Fisher’s exact, χ^2^, Wilcoxon and regression analyses are shown in full in **Supplementary Tables 4 & 5**.

Second, in germline heterozygotes (group 2), there was no tendency for the wildtype copy of *MUTYH* to be lost over the mutant (OR=0.530, 95%CI 0.026-8.30, P=1.00, Fisher’s exact). The very small number of mono-allelic *MUTYH* carriers with somatic loss of the wildtype allele in their tumours (group 2a, n=2) limited the comparison with groups 2b/2c, although C:G>A:T activity was a little higher in the former (**Figure 3**). There was also evidence that the resulting MUTYH deficiency in the group 2a CRCs could specifically cause SBS36 rather than SBS18 (**Supplementary Figure 1**). No mutations were assigned to SBS36 in mono-allelic *MUTYH* carriers that did not have loss of the wildtype allele (groups 2b/2c, n=42).

Third, germline heterozygotes as a whole (group 2) had higher SBS18 metrics and C:G>A:T activity than *MUTYH* wildtype cases (group 3). Half of the 44 individuals in group 2 exhibited SBS18 in their CRC, representing a highly significant increase over group 3 (2.7-fold odds ratio, *P=*4.48x10^-4^; **Table 1**, **Figure 4**, **Supplementary Tables 1 & 2**). This association remained strong (2.7-fold odds ratio, P=3.00x10^-3^) when the two group 2a CRCs were excluded, indicating that it was not driven by somatic loss of the wildtype *MUTYH* allele. SBS18 mean burden and activity were both about 2.5-fold higher in group 2 than in group 3 (**Figure 4**). In comparison, SBS36 metrics were similarly low in groups 2 and 3. In multivariable analysis (details not shown), significantly higher SBS18 and C:G>A:T metrics were independently associated with group 2 (*versus* group 3), absence of MMRd and, like SBS18+ *MUTYH-*wildtype CRCs [21], cancer location in the proximal colorectum.

**Figure 4.**
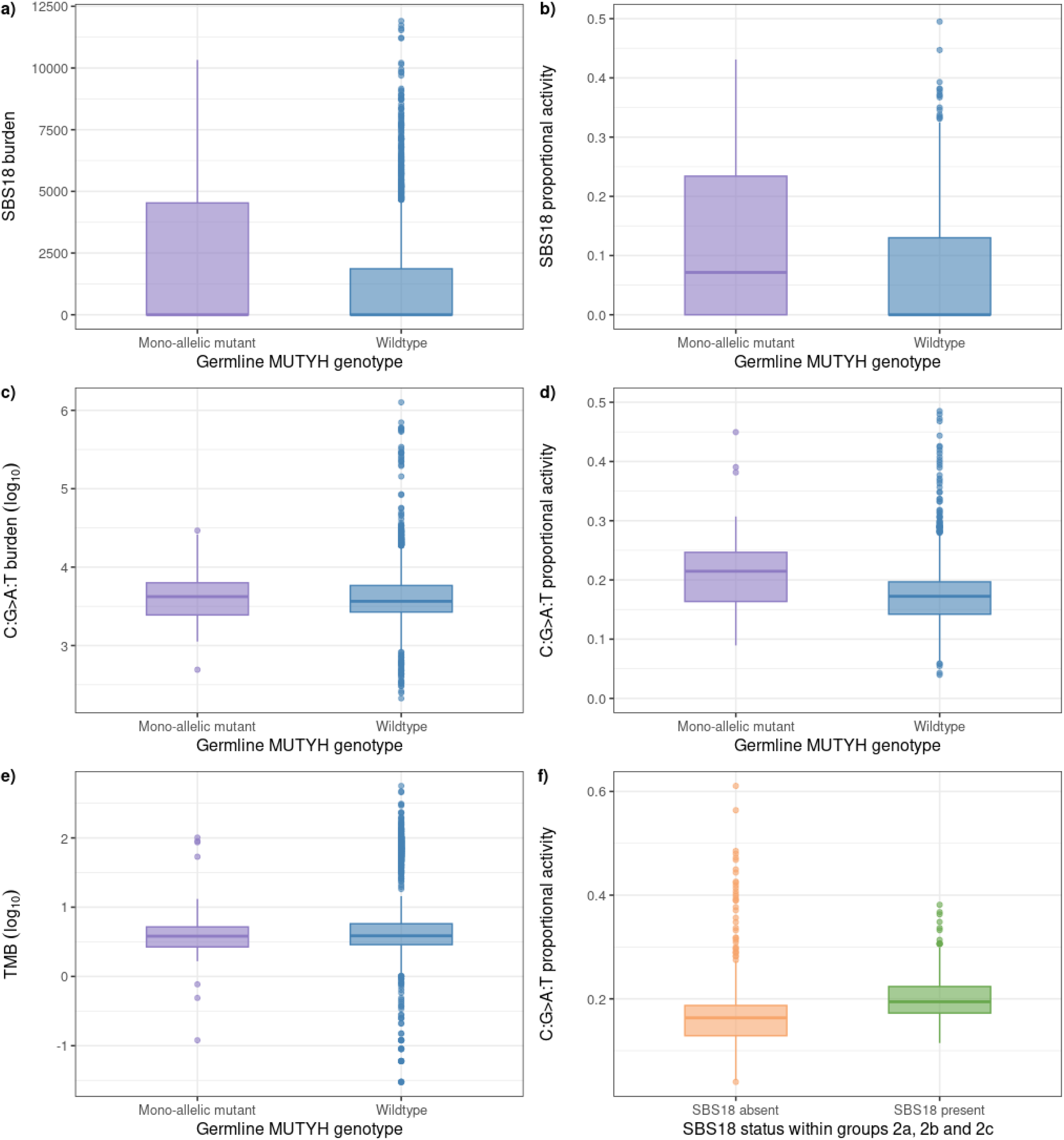
Associations in all 100kGP CRCs between measures of somatic mutation and germline heterozygote MUTYH status (group 2) versus vs wildtype (group 3). (a) SBS18 burden; (b) SBS18 activity; (c) C:G>A:T burden; (d) C:G>A:T activity; and (e) TMB (measured as somatic coding variants per Mb). Panel (f) shows the expected positive relationship between the presence of SBS18 and C:G>A:T activity within group 2; a weaker positive association was seen with C:G>A:T burden (not shown). Log scales are used to aid display in some cases. Nominally significant associations (*P*<0.05) were present based on Wilcoxon tests for panels (a), (b) and (d). Associations from Fisher’s exact, χ^2^, Wilcoxon and regression analyses are shown in **Table 1** and **Supplementary Tables 3**-**5**.

To avoid associations between mono-allelic *MUTYH* mutations and tumour features being confounded or obscured by the approximately six-fold increased SBS burden of MMRd cancers that is biased to C:G>T:A changes [23], we repeated the above analyses in MSS/polymerase proofreading-proficient CRCs only (37 group 2 and 1,875 group 3). The associations between group 2 and increased SBS18 and C:G>A:T were generally very similar to those in the all-CRC analyses (**Supplementary Tables 1-8**). Of note, however, in MSS-only regression analyses corrected for the number of non-C:G>A:T mutations in each cancer, TMB was significantly increased in mono-allelic *MUTYH* carriers (group 2) compared with the *MUTYH*-wildtype individuals of group 3 (for SCVPM measure of TMB, β=0.447, SE 0.223, P=0.045; for total SBS burden, β=1839, SE 725, P=0.011). The equivalent association was not detected in an MMR-proficient-only analysis, perhaps unsurprisingly given the much higher background level of SBSs in those tumours (for SCVPM, β=-10.3, SE 5.99, P=0.086; for total SBS burden, β=1160, SE 1623, P=0.475). Mono-allelic *MUTYH* carriers contributed 4.1% of all SBS18 mutations in MSS CRCs cancers (**Supplementary Table 2**).

Given that MAP patients’ colorectal tumours are known to acquire extra C:G>A:T mutations in driver genes owing to bi-allelic MUTYH deficiency [13], we tested whether the CRCs of mono-allelic mutation carriers (group 2) also tended to acquire C:G>A:T driver changes. This would be expected if the associated increased SBS18 activity occurred prior to, or early in, colorectal tumorigenesis. Based on a pre-specified analysis of the 20 most common CRC driver genes [21], we found that group 2 patients’ CRCs did indeed carry an increased proportion of C:G>A:T driver changes (**Table 2**).

**Table 2.** Associations between driver mutation spectra in the 20 most frequently mutated CRC driver genes and mono-allelic MUTYH mutations. Analysis restricted to MSS cancers. OR=1.708, 95%CI 1.046-2.79; P=0.0306, χ^2^1 test.

| Driver gene mutation | *MUTYH* mono-allelic | *MUTYH* wildtype | Total |
| --- | --- | --- | --- |
| C:G>A:T | 23 | 908 | 931 |
| Other SBS | 56 | 3776 | 3832 |
| Total | 79 | 4684 | 4763 |

In an analysis focussed solely on SBS18-positive CRCs, we investigated whether both the C:G>A:T mutations and other somatic molecular features of the cancers were similar in groups 2 and 3, so as to identify any distinctly different mechanisms of mutagenesis in the two groups. SBS18 burden/activity and C>A burden/activity were all significantly higher in group 2 in univariable analysis, broadly consistent with the all-CRC analysis (**Supplementary Table 8**), but the two groups were similar in terms of SCVPM, age, location, whole genome doubling and MMRd (**Supplementary Tables 1-8**). These results were confirmed in multivariable analyses (not shown). They were consistent with a simple model in which both groups are exposed to similar levels of oxidative DNA damage, but damage repair is less effective in group 2 patients.

### Associations between germline mono-allelic MUTYH mutations, SBS18 and C:G>A:T mutations are also present in CRCs from the Hartwig collection

We tested whether the associations between mono-allelic *MUTYH* mutations and mutational processes in the 100kGP CRCs were present in an independent data set. We derived germline *MUTYH* genotypes and somatic mutation data from 493 metastatic CRCs in the Hartwig study [24, 25]. Although many of the tumours analysed had been subjected to genotoxic therapy, we still observed significantly higher SBS18 burden, SBS18 activity and C:G>A:T burden in the 14 CRCs from group 2 compared to the group 3 patients (**Supplementary Table 9**).

### Association study of CRC risk in carriers of germline MUTYH mutations and causality analysis

In order to explore the possibility that increased SBS18 activity and TMB led to a raised risk of CRC in germline *MUTYH* heterozygotes, we utilised three sets of CRC cases and controls: (i) UK 100,000 Genomes Project (100kGP, https://doi.org/10.6084/m9.figshare.4530893.v7) supplemented with our own CORGI study; (ii) UK Biobank exomes; and (iii) UK Biobank genomes [26] (**Figure 5**). We filtered participants to ensure ancestral matching and exclusion of multiple close relatives. We also excluded individuals with probable bi-allelic germline *MUTYH* mutations. Common *MUTYH* polymorphisms (allele frequency >0.025) and other variants not predicted to be pathogenic from ClinVar (https://www.ncbi.nlm.nih.gov/clinvar/?term=%22mutyh%22[GENE]) were also excluded. Meta-analysis of the cohorts, comprising 4,834 cases and 138,005 controls, demonstrated an association between *MUTYH* heterozygote status and CRC (OR=1.204, 95%CI 1.044-1.388; *P=*0.0108; **Figure 5**, **Supplementary Table 10**).

**Figure 5.**
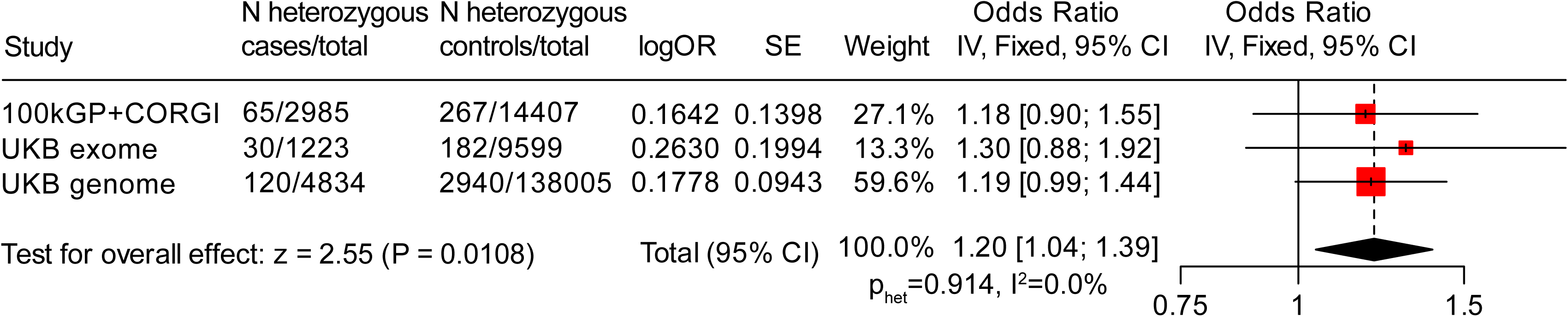
Inverse variance-weighted meta-analysis of association between CRC/polyposis and MUTYH heterozygote status (cases) in 100kGP, UKB and Exome array GWAS. Black dots show point estimates of odds ratio (OR) for each study, with whiskers showing 95%CIs. Squares are proportional to study size. The red line shows the pooled OR and the diamond the pooled 95%CIs. There was no evidence of inter-study heterogeneity. See also **Supplementary Table 10**.

Since about half the CRCs from germline *MUTYH* heterozygotes did not have SBS18, we examined candidate modifier SNPs for negative effects on *MUTYH* expression that might help to ‘reveal’ haploinsufficiency in heterozygotes’ CRCs [27]. The SNP set was restricted to a ∼2Mb window flanking *MUTYH* (hg38 chr1:44,329,242-46,339,970) and to common SNPs from dbSNP155 (https://genome-euro.ucsc.edu/cgi-bin/hgTrackUi?hgsid=407686972_tMiyAfTray8NrAweTgTfYMqHxAx6&db=hg38&c=chr1&g=dbSnp155Composite), and included several strong expression- and splice-quantitative trait loci for *MUTYH* (https://www.gtexportal.org/home/gene/MUTYH). However, we found no evidence of SNP alleles associated with the presence of SBS18 in mono-allelic *MUTYH* carriers (uncorrected *P>*0.05 in all cases, details not shown). Functional *MUTYH* haploinsufficiency, perhaps coupled with environmental exposure to oxidative damage, thus seems the most likely causal mechanism.

The principles of Mendelian randomisation [28, 29] indicated that mono-allelic germline *MUTYH* mutations are probably causal for increased SBS18, and hence that SBS18 may be causal for CRC. A causal link between SBS18 and raised CRC risk was supported by a Wald ratio estimation for the SBS18-CRC association (*mrratio* command in STATA, β_SBS18-CRC_=0.177, SE 0.085, OR=1.19, *P=*0.037) [30]. We also performed a comprehensive check for sources of collider bias and other confounders that could have led to a spurious association between mono-allelic *MUTYH* mutations and SBS18 in CRC, including factors such as genetic ancestry and geographical location that could in principle co-vary with germline genotype, but no evidence of any such confounder was found (**Supplementary Tables 11 & 12)**.

### Mutational features in germline MUTYH heterozygotes in the 100kGP all-cancer cohort

The findings from CRC suggested that the effects of *MUTYH* heterozygosity might be pervasive, in that germline heterozygous genotypes could lead C:G>A:T mutations to accumulate in many normal and/or tumour cell types. We therefore examined SBS18, SBS36 and C:G>A:T mutations in the 100kGP All-cancer data set, which comprised 15,223 patients and tumours from multiple tissue locations. Owing to the inherent variation among these patients, we took a conservative approach and restricted this part of the study to assessing the three major findings from CRC (that is, higher SBS36 in group 1 than 2&3, occasional SBS36 in group 2a compared with 2b/c, and higher SBS18 in group 2 than 3).

We here present data from analyses of all cancer types and of all cancers except CRC (‘CRC-excluded’) (**Table 1**, **Supplementary Tables 13-16 and Supplementary Figure 3**).

We first noted that CRCs generally had much higher SBS18 (and somewhat higher SBS36) metrics than other cancer types (**Figure 3**, **Supplementary Tables 1**, **2** & **13**-**16**), including approximately three-fold higher SBS18 prevalence, but 10-fold higher SBS18 activity and C:G>A>T burden. In the germline *MUTYH* heterozygotes (group 2), we identified 25 non-CRC cancers with LOH of the wildtype or mutant *MUTYH* allele (groups 2a and 2b respectively). As in CRC, there was no tendency for the wildtype copy of *MUTYH* to be lost over the mutant (OR=0.778, 95%CI 0.220-2.786, P=0.778, Fisher’s exact). Furthermore, the 10 cancers in which the wildtype allele was lost (group 2a) were from eight different tissue or cell types, providing no evidence that loss of the wildtype was provided a selective advantage in specific tumour types. No cancer had bi-allelic somatic *MUTYH* mutations (including LOH and homozygous deletions).

Overall, despite the multiplicity of tumour types in the CRC-excluded analysis and the less frequent C:G>A:T changes, the CRC-excluded data strongly supported the findings from CRC on C:G>A:T mutations and mutational signatures (**Table 1, Supplementary Tables 14-17**). An excess of SBS36 and SBS18 was present in bi-allelic *MUTYH* mutation carriers’ tumours (group 1 *vs* 2&3). SBS36 was increased in the tumours of heterozygotes that acquired somatic loss of the germline wildtype allele (group 2a *vs* 2b/c), with accompanying increased C:G>A:T activity. There was particular support for the association between heterozygous germline *MUTYH* mutations and SBS18 (group 2 *vs* 3), with effect sizes (as measured by odds ratios) similar to the CRC-only analyses (**Table 1**). In fact, mono-allelic *MUTYH* carriers contributed 8.2% of all SBS18 mutations in CRC-excluded cancers, a significantly higher proportion than in MSS CRCs (*P<*10^-7^, **Supplementary Tables 1, 2, 15 & 16**).

We performed specific analyses of cancers of the upper gastrointestinal tract (sometimes found in MAP), breast (common female-predominant sporadic cancer), prostate (common male-predominant sporadic cancer) and endometrium (major part of Lynch syndrome and polymerase proofreading polyposis). The analyses were limited by smaller sample sizes and/or lower SBS18 metrics than CRC and All-cancer, but provided good support for higher SBS18 (or SBS36) and/or raised C:G>A:T levels in (i) group 2a *vs* 2b/c and (ii) group 2 *vs* 3 (**Supplementary Tables 16, 18 & 19**).

In summary, despite the many different tumour types in the All-cancer and CRC-excluded analyses, the positive effects of bi-allelic and mono-allelic germline *MUTYH* mutations on mutational processes (SBS36, SBS18 and C:G>A:T changes) were clearly present, as they were in CRC. Given the much higher baseline SBS18 and C:G>A:T metrics in CRCs than other cancers, the importance of germline heterozygous *MUTYH* mutations, and the resulting C:G>A:T changes and SBS18 may be much greater as a driving force of tumorigenesis in CRC. Nevertheless, the SBS18, SBS36 and C:G>A:T data suggested that some increased risk of other cancers in bi-allelic and heterozygous *MUTYH* mutation carriers might exist.

### The risks of multiple cancer types in carriers of heterozygous germline MUTYH mutations

One of the purposes of the current study was to determine whether the associations between *MUTYH* genotype and cancer risk reported by others, such as the multi-cancer analysis of Barreiro et al [18], were also present in the large UK cancer data sets. After including only unrelated individuals of European ancestry and excluding CRC or polyp cases, we identified 13,252 cases with any cancer and 14,407 cancer-free controls in the 100kGP, plus 14,813 cases and 9,599 controls from UKB. Bi-allelic *MUTYH* mutations were associated with no cancers other than CRC (**Supplementary Tables 20-23**).

For assessing associations between All-cancer (CRC-excluded) risk and *MUTYH* germline heterozygosity, there was estimated 70% power to detect an association at nominal *P=*0.05 if *MUTYH* heterozygosity conferred a relative risk of 1.20 (https://csg.sph.umich.edu/abecasis/gas_power_calculator/index.html). However, we found no association between heterozygous germline *MUTYH* mutation and CRC-excluded cancer risk in a meta-analysis of 100kGP and UKB data (OR=0.93, 95%CI 0.82-1.06, *P_meta_=*0.58).

Since there have been variable reports that *MUTYH* heterozygosity conferred risks of cancers of specific organs other than CRC [7, 17, 18, 31–38], we also assessed these in 100kGP and UKB. No cancer type achieved *P<*0.10*_meta_* (**Supplementary Tables 24 & 25**), reflecting limited power.

### De novo mutations in the offspring of MUTYH heterozygotes

We searched for effects of *MUTYH* heterozygosity on germline *de novo* mutation (DNM) rates in parent-offspring trios from 100kGP [39]. We found 206 trios in which one parent was a *MUTYH* heterozygote. Compared with 11,308 *MUTYH* wildtype control trios, there was a small, but significantly increased total *de novo* mutation burden in the offspring of *MUTYH* carrier parents assessed using *standard criteria* (median 948 v 915, *P=*0.016), but not using *stringent criteria* (median 69 v 70, *P=*0.55), which is the measure recommended by 100kGP (see **Methods** for link to definitions). We also specifically assessed *de novo* C:G>A:T mutations and found no association (median 6 for both groups, P=0.783, stringent criteria).

We concluded that any effect of mono-allelic *MUTYH* mutations on DNMs was likely to be small at most (**Supplementary Table 24**).

### Other base excision repair heterozygotes may be predisposed to specific mutational signatures

We explored whether the association between germline heterozygotes, somatic mutations and cancer risk was a special feature of *MUTYH*, or was present for other BER genes.

*MBD4* was chosen for study, because its recessive syndrome is genetically similar to MAP and includes multiple polyps and CRC, and no heterozygote risk of colorectal tumours has been established [40–43]. Bi-allelic germline *MBD4* mutations predispose to increased CpG>TpG changes, resulting in an SBS1-like signature. Apart from a raised (yet very small absolute) risk of uveal melanoma in *MBD4* heterozygotes owing to somatic loss of the germline wildtype allele [44], no increased cancer risk in *MBD4* heterozygotes is known.

We found the frequency of LoF germline mutations in *MBD4* to be ∼0.1%, over an order of magnitude lower than that for *MUTYH.* In 100kGP All-cancer, after excluding two cancers with second hits, 14 *MBD4* germline mutation carriers were present. Compared with over 14,000 All-cancer *MBD4-*wildtype cases, *MBD4* germline heterozygosity was associated with higher SBS1 burden, SBS1 activity and C:G>T:A activity in tumours at nominal *P<*0.05 in univariable analysis. SBS1 burden (*P=*0.0143), SBS1 activity (*P=*0.0096) and C:G>T:A burden (*P=*0.00201) remained significantly associated in multivariable analysis (**Supplementary Table 25a**). In CRC-excluded cases, mono-allelic *MBD4* status was associated in univariable analysis with SBS1 burden and activity, whereas associations in the smaller CRC data set did not reach statistical significance. Given the evidence that *MBD4* heterozygotes can cause an excess of C:G>T:A mutations, we revisited the question of whether such individuals are predisposed to CRC in 100kGP+CORGI and UKB. Whilst further studies are required, we found an increased risk of CRC in the mono-allelic *MBD4* mutation carriers (OR=1.75, *P=*0.020; **Supplementary Table 25b**).

## Discussion

Our study is one of the biggest yet to consider cancer risk in carriers of germline mono-allelic *MUTYH* mutations. Our results suggest that these individuals do have a moderately (∼1.2-fold) increased risk of CRC. We took steps to exclude bias in the analysis, to the greatest degree possible. We specifically believe it unlikely that cryptic second germline *MUTYH* mutations in the mono-allelic mutation carriers (group 2) can explain the association we find between mono-allelic *MUTYH* and SBS18 – this is partly because we used WGS data and partly because SBS36 was highly specific to cancers with bi-allelic germline *MUTYH* mutations or somatic loss of the wildtype allele (group 2a). No group 2b/c CRCs (without somatic loss of the wildtype allele), and only 3/253 group 2 non-CRCs, showed SBS36, despite its similarity to SBS18 and thus hypothetical possibility of chance mis-assignment (**Supplementary Tables 1, 2, 14 & 15**). Our association study did not support previously reported increased risks of extra-colonic cancers in carriers of mono-allelic or bi-allelic germline *MUTYH* variants, although very small effects remain possible.

Other groups have proposed that somatic loss of the wildtype *MUTYH* allele causes increased cancer risk in germline *MUTYH* heterozygotes. Our findings differ considerably from those studies. For example, the largest, most recent such study of multiple cancer types [15] used molecular and bioinformatic methods distinct from ours to study FFPE samples from >350,000 Foundation Medicine tumours, enriched for metastatic disease, analysed on a 311- or 324-gene diagnostic panel. Constitutional DNA was absent, and hence *MUTYH* variants were assigned as germline or somatic using an algorithm with stated accuracy 80-85% [45]. Somatic copy number data were estimated from the sequence data, although copy-neutral loss-of-heterozygosity could not be assessed and heuristic modifications of uncertain types were made to the genotype calling. The authors reported loss of the germline wildtype allele in 12% of mono-allelic mutation carriers (our group 2a), but no tumours had loss of the mutant allele (our group 2b). Thirty SBS signatures (COSMIC version 2) were derived from the 324-gene data using a method developed solely for hypermutant tumours (>13.8 mutations/Mb) and thus potentially unsuited to the majority of cancers [46]. An SBS18 score of unknown origins was assigned, although this may also have detected SBS36. No controls were analysed and no driver gene somatic mutation data were provided. It is hard fully to assess these data, when, for example, algorithmic mis-assignment of a few bi-allelic germline mutations could profoundly influence the conclusions. Nevertheless, we contend that our study has its own strengths, especially the availability in 100kGP of paired constitutional DNA and whole-genome sequence data from fresh-frozen tumour samples, collected, prepared and analysed using a single pipeline. We suggest that our data are likely to provide accurate identification of second hits at *MUTYH*, assignment of mutational signatures, and calling of CRC driver gene mutations. More generally, the 100kGP data show the benefits of tumour genomics in determining mechanisms of increased CRC, risk by allowing the full assessment of mutational spectra and signatures that can act as causal mediators.

Our data do show that somatic LOH of the wildtype allele can occur in the tumours of a small minority of mono-allelic *MUTYH* carriers (group 2a), and that this sometimes elicits the MAP signature SBS36. However, whilst somatic loss of the wildtype *MUTYH* allele in ∼5% of mono-allelic mutation carriers could in theory explain the estimated increased risk of CRC in group 2 as a whole, the ‘second hit’ mutation should be selected and thus much more common than loss of the mutant allele (i.e. group 2a should be much larger than group 2b). We found no evidence of this. Furthermore, about 90% of mono-allelic carriers showed no somatic loss in their CRCs (group 2c), yet still exhibited the associations with SBS18. Our evidence thus indicates that haploinsufficiency, acting through raised SBS18 and hence increased TMB and driver gene mutations, is the main mechanism of increased CRC risk in all mono-allelic *MUTYH* carriers. A plausible scenario is that exposure to extrinsic or intrinsic oxidative stress periodically overwhelms the capacity of a single functional *MUTYH* allele to repair the resulting DNA damage. In our opinion, much of the SBS18-associated DNA damage in tumours from carriers of mono-allelic germline *MUTYH* mutations (group 2) may occur in precursor lesions or even normal cells prior to being ‘revealed’ in the clonally expended cancer cells. This can explain the similar SBS18 levels in cancers with and without second hits at *MUTYH,* and the lack of excess SBS18 in CRCs with somatic mono-allelic deletion of one copy of *MUTYH* and no germline mutation. The evidence that somatic loss of *MUTYH* is not advantageous for tumorigenesis appears to conflict with the evidence that moderate hypermutation in *MUTYH* germline heterozygotes increases tumour risk.

However, this may simply reflect the near-ubiquitous effects of germline alleles that are selected at the level of the organism, whereas somatic mutations in ‘caretaker genes’ have to survive strong competition from thousands or millions of other cells in the same tumour, even at the pre-malignant stage [47, 48].

The detection of increased SBS18 and C:G>A:T somatic mutations in the All-cancer and CRC-excluded tumour sets was both important and not fully expected. Of note, however, Moore *et al* [49] found SBS18 to be active in 19/29 normal cell types, consistent with our findings. The lower baseline SBS18 levels in the CRC-excluded tumours may limit the effects on cancer risk, consistent with our failure to detect raised risk of extra-colonic malignancies in our data [7, 18, 33, 35]. We also found no evidence of increased *de novo* mutations in *MUTYH* heterozygotes, again consistent with Moore *et al* [49] who did not detect SBS18 in seminiferous tubules.

Associations have variably been reported previously between an activating germline *APOBEC3A/B* deletion and APOBEC-associated hypermutation [50–56]. This leads to increased breast cancer risk, with a similar effect size to that of *MUTYH* heterozygotes on CRC. However, APOBEC is an anti-viral, pro-mutational process, as distinct from the DNA repair deficiency in our *MUTYH* and *MBD4* mutation carriers. The *APOBEC3A/B* deletion also only seems to have its effects on mutational signatures and tumour risk in breast cancers, whereas we find our *MUTYH* and *MBD4* effects in pan-cancer analyses and probably in many normal tissues.

In conclusion, *MUTYH* heterozygotes have increased risk of CRC owing to haploinsufficiency, increased C:G>A:T mutations and raised activity of signature SBS18. We have shown more generally that non-rare germline variation in DNA repair genes can lead to specific mutational signatures and hypermutation in multiple tumour types, without somatic second hits being required. The magnitude of increased CRC risk is moderate in *MUTYH* heterozygotes – although higher than most common risk polymorphisms – and it is possible that some such individuals exposed to high levels of reactive oxygen species [57] are at more elevated risk. It is plausible that *MBD4* heterozygotes also have an increased risk of CRC through similar mechanisms, although the rarity of these individuals mandates some caution in this regard. In total, we estimate that about 5% of the population could be germline heterozygotes for apparently recessive DNA repair genes. Further large studies will be required to confirm this estimate and explore the clinical utility of identifying such individuals. Whilst the *MUTYH* (or *MBD4*) alleles appear to constitute a valid instrumental variable for CRC risk, acting through the intermediate phenotype of SBS18 (or SBS1), it seems that the net effect on risk is absent or undetectable for many cancer types. It is arguably reassuring that a two- to three-fold increase in the activity of a common signature, like SBS18 in CRC, has a 1.2-fold effect on cancer risk, and that effects on the risk of other cancers are undetectable. Nonetheless, we contend that polygenic risk scores for CRC should include *MUTYH* genotypes. Furthermore, despite past controversies [58], measures to reduce oxidative damage exposure could indeed be an effective CRC prevention strategy in general, as long as suitable agents can be developed.

## Methods

### 100kGP and CORGI CRC and multiple polyp cases

Cases were individuals with colorectal carcinoma and/or multiple colorectal polyps who were recruited to (i) the 100kGP colorectal cancer domain owing to resection of biopsy of a colorectal carcinoma, (ii) the 100kGP familial colorectal cancer or multiple polyp domain, or (iii) the CORGI study (familial, early-onset or multiple CRCs and/or polyps). An individual maximum pairwise kinship coefficient threshold of 0.01625 was applied and individuals had predicted European ancestry >0.99, leaving 2,985 eligible cases. For each case, WGS data from constitutional DNA to median depth >30X were obtained using the Illumina Hi-Seq platform, together (in category (i)) with paired frozen cancer sample WGS to median depth >100X in many cases.

### 100kGP controls

Data included in release version 14 of the rare disease domain in the 100kGP were used to create the control cohort of individuals. Included individuals had no clinical record of cancer (of any site), or a condition closely linked to cancer or malignancy (including not only colorectal polyp(s), but also other benign tumours or clonal haematopoiesis). Ancestry and kinship filters were applied as for 100kGP cases, leaving 14,407 eligible individuals with WGS data. Whilst 100kGP controls were younger than cases (median age 43 *versus* 68 respectively for CRC), we regarded the controls as representative of *MUTYH* allele frequencies in the European ancestry population and the inclusion of younger individuals was in any event conservative.

### UKB cases

WES data were obtained under UK Biobank project ID 19655 and WGS data were obtained under project ID 155012. All individuals with a current diagnosis or personal history of colorectal cancer were identified based on International Classification of Diseases ICD-10 codes C180, C182-189, C19 or C20 using data field 40006 or ICD-9 codes 1530-1534 or 1536-1541 using data field 40013. We further included only individuals with tumour behaviour described as "malignant, primary site" in data field 40012 and with histology information available in data field 40011 to confirm the CRC diagnosis. Cancers which were self-reported only (provided in data field 20001) could not be validated and were therefore excluded. Patients with colorectal polyps only were excluded, due to a lack of details on the polyp number and size. We then applied genetic ancestry and kinship filters to select for Europeans and unrelated individuals using the same threshold values as the other cohorts (>0.99 and <0.01625 respectively), leaving 1,224 cases with WES data and 4,834 non-overlapping cases with WGS data.

### UKB controls

From approximately 200,000 participants with WES available, we excluded any individuals with an ICD-9 or ICD-10 cancer, or self-reported cancer diagnosis (provided in data fields 40013, 40006 and 20001 respectively), or reports of colorectal polyp(s) based on ICD-10 codes D12, D120, D121, D122, D123, D124, D125, D126, D127, D128 or D129) in data fields 41202 and 41204, or ICD9 codes 211, 2111, 2112, 2113, 2114, 2115, 2116, 2117, 2118 or 2119 in data fields 41203 and 41205, or code 1460 in data field 20002. We also removed all individuals with reported history of cancer in either parent or any sibling of the cancer types with data available (breast, lung, bowel or prostate cancers). We then applied genetic ancestry and kinship filters as above. As the potential cohort size for the controls (N=108,841) was far in excess of cases, we randomly sampled 10,000 and 30,000 individuals to form the control cohorts for CRC/polyposis and All-cancer analyses respectively. The ages of UKB WES controls were similar to those of cases (medians 56 *versus* 63 respectively). For the UKB WGS control cohort, we extracted 138,005 non-overlapping cancer-free individuals using the same filters and thresholds.

### 100kGP and UK All-cancer case cohorts

From all unrelated individuals from the 100kGP All-cancer domain with European genetic ancestry, we selected 11,318 participants with a diagnosis of a malignant neoplasm described by ICD-O-3 in their clinical history. This All-cancer cohort used individuals only from the 100kGP (no additional WGS data was included) and did not contain participants with only colorectal polyposis. From UKB, we selected participants with a malignant neoplasm using ICD9 and ICD-10 data fields 40013 and 40006 respectively, and we then converted these into ICD-O-3 codes. After genetic ancestry and kinship filters, 12,694 UKB cases remained. In both 100kGP and UKBioBank, we grouped cancers into 20 organ sites using the SEER recode paradigm (National Cancer Institute, 2008), whilst keeping colorectal cancers, upper gastrointestinal tumours, and cancers of the liver and intrahepatic bile duct as three separate categories for our analyses. We also selected for more specific tumour subtypes of interest. In 100kGP, this was performed by matching for the appropriate specific terms for tumour site and organ of origin in histological and other clinical records. In UKB, we extracted patient information with the following ICD codes, in order to facilitate comparisons with previous studies:

- Adrenocortical carcinoma (749 in ICD9; C740 in ICD10)
- Oesophageal carcinoma (1505 or 1509 in ICD9; C150-C159 in ICD10)
- Prostate adenocarcinoma (1850 in ICD9; C61 in ICD10)
- Renal clear cell (1890 in ICD9; C64 in ICD10)
- Sarcoma (1710-1719 in ICD9; C490-C499 in ICD10)
- Acute myeloid leukaemia (AML) (2050 in ICD9; C920 in ICD10)
- Uveal melanoma (1900, 1905, 1906 or 1909 in ICD9; C693 or C694 in ICD10)

### Identifying deleterious germline variants in MUTYH

For 100kGP data, germline variants called and filtered by Starling (v2.4.7) within the Illumina WGS workflow were already available in the 100kGP research environment using the GRCh38 reference genome [59]. For UKB, germline variant calls and filtering information from DeepVariant (v0.10.0) were already available [60]. For both GEL and 100kGP, we selected all variants that passed the variant calling filtering criteria and intersected with the chromosomal location of *MUTYH*. We then re-annotated all variants using Ensembl VEP (v109.3) [61] to obtain the full predicted functional consequences. We first filtered in PTVs, and then added missense and any other variants annotated as pathogenic or likely pathogenic in ClinVar [62]. Whilst a small number of variants had conflicting interpretations of pathogenicity in ClinVar, closer expectation showed that the great majority of these were most likely to be pathogenic and only a single submission was of uncertain significance.

Variants predicted by SpliceAI to have a splice donor or acceptor functional probability score > 0.8 were annotated as pathogenic, as the recommended threshold [63]. Germline structural variants were search for using MANTA. Individuals homozygous for a predicted (likely) pathogenic variant were classified as having bi-allelic *MUTYH* mutations, as were those with two different predicted (likely) pathogenic variants. Individuals with a single pathogenic germline variant of heterozygous genotype were classified as having mono-allelic loss of gene function, and the remainder of the cohort were categorised as germline wildtype for *MUTYH*.

### Somatic mutation analyses in 100kGP tumours

To investigate how germline mutations in *MUTYH* influence tumorigenesis in colorectal cancer and other cancer types, we identified patients with somatic sequencing data in 100kGP. This was not restricted to unrelated participants or those with European genetic ancestry. For ‘second hits’ and other somatic mutations at *MUTYH*, including somatic copy number changes and copy-neutral LOH (cnLOH), we used Battenberg (v2.2.8) [64], where these data were available, or ASCAT (v3.0.0) [65]. Read depths of the reference and alternative (*MUTYH* mutant) alleles were also assessed in tumour (after correcting for purity) and normal sequence, particularly for determining whether the mutant or wildtype copy of the *MUTYH* gene was lost in tumours with LOH. LOH was scored when one copy of the *MUTYH* locus was absent in the cancer, with cnLOH (zero copies of one allele) if the other chromosome was also at the modal number in that tumour. We detected no homozygous deletions or high-level amplifications, and thus all other copy number states were scored as ‘no loss’ (NL). We also searched for pathogenic somatic mutations in *MUTYH* using existing somatic variant results from Strelka (v2.4.7) .

### Identifying additional DNA repair deficiencies in cancer genomes

Microsatellite instability (MSI) was assessed using mSINGS [66] as a proxy for MMRd. The multiplier parameter (the number of standard deviations away from the baseline required to call instability) was configured to 2.0. The maximum fraction of unstable sites permitted in order to call a cancer MSI-negative was set to 0.2, and the minimum proportion of unstable sites to call a sample MSI+ was also 0.2. The selection of the sites used and the success of the algorithm in distinguishing MSI+ and MSI-negative CRCs in the 100kGP data set has been described previously [21]. We also identified all tumours with known pathogenic germline or somatic mutations in the DNA polymerase genes *POLE* or *POLD1* [67]; these were classified as polymerase-proofreading (POL)-deficient.

### Mutational spectra and signature analysis

Tumour mutation burden (TMB) for each of the tumour samples was defined as the total number of pass-filtered non-synonymous SNVs and indels shorter than 50 base pairs, divided by the total length of the coding sequence (32.61 Mb) (see https://re-docs.genomicsengland.co.uk/cancer_clinical/). We extracted all single base substitution (SBS) mutations present in each tumour in their trinucleotide (96-channel) context using the tool SigProfilerMatrixGenerator (v1.2.1) [68] and then inputted the data to SigProfilerAssigment (v1.2.1) [69] in order to assign corresponding mutational signatures from the COSMIC (v3.3) reference database. SigProfiler was implemented within Python v3.6.8 using GRCh38 as the reference genome and all other parameters at the recommended default values. Mutations in each channel or signature were generally assessed in three ways: (a) *prevalence* (presence at any level >0% or absence) in a cancer; (b) *burden* (absolute number of a mutation type in a cancer); and (c) *proportional activity* (the total burden of a defined class of mutation relative to all mutations of that type). Examples of measures of proportional activity (sometimes referred to simply as ‘activity) included C:G>A:T mutations relative to all SBSs or mutations assigned to signature SBS18 relative to all mutations assigned to a SBS signature. The three mutational measures co-vary, but each has specific strengths. Prevalence is non-quantitative, but is useful for determining whether a specific signature (and underlying mutational process) is active within a tumour; it appears to be a relatively sensitive and specific measure (e.g. despite its resemblance to SBS18, SBS36 is only very rarely found outside the context of MUTYH deficiency). Burdens are particularly useful for identifying hypermutant tumours, owing to genomic instability, environmental exposure or unknown factors. Proportional activity is useful when correcting for background burdens to identify the most active processes in a single tumour; one use is to identify important mutational processes that are not found by crude TMB measures owing to ‘competing’ mutational processes (*e.g.* in a process of parallel evolution, different CRCs may derive the mutations they require from different sources without major effects on TMB).

### Colorectal cancer driver gene analysis

To examine the effects of somatic *MUTYH* genotypes on colorectal tumorigenesis, we extracted driver mutations from the following 20, relatively common established CRC driver genes in 100kGP version 19 (*ARID1A, BCL9, ZFP36L2, TGFBR2, PIK3CA, FBXW7, APC, BRAF, KMT2C, PTEN, TCF7L2, ATM, BCL9L, KRAS, TP53, RNF43, SOX9, SMAD4* and *ASXL1*) [21]. Specifically, we extracted small mutations (almost all SBS or indel) and assigned pathogenicity, including bi-allelic mutations for tumour suppressor genes.

### De novo mutations

WGS data [39] were available from 13,912 trios (both parents and a child) within the 100kGP project, accompanied by sets of *de novo* mutations in the child that were present in neither parent and were presumed to have arisen in the parental gametes, or perhaps at an early stage of the child’s development. We assessed *de novo* mutation burdens in these trios according to the parental *MUTYH* variant status using data mapped to genome build 38.

Further details of the 100kGP *de novo* mutations project and their recommendations for use of the *stringent m*utation sets can be found at https://re-docs.genomicsengland.co.uk/de_novo_data/.

### Germline MBD4 mono-allelic mutation carriers

Analysis was generally performed as for *MUTYH.* We excluded MMRd tumours to avoid background somatic mutations that are known to occur, sometimes as sub-clonal events, within a short coding repeat in *MBD4*. One mono-allelic mutation carrier with a second hit by LOH in their tumour was also excluded from the analysis of associations with somatic mutational features. Seventeen uveal melanoma cases were identified and excluded, although none were heterozygous for *MBD4*. No bi-allelic *MBD4* mutation carriers were found. SBS1 burden, SBS1 activity, C>T burden, C>T activity and SCVPM were used as outcomes to assess the effects of germline mono-allelic *MBD4* mutations in All-cancer analyses only, owing to limited power in CRC-only analyses.

### Statistical analysis

All statistical analyses were performed in R (v4.2.1). All figures were produced using the R package ggplot2. Germline analyses used multiple logistic regression for the binary case-control outcome. Meta-analyses were performed using fixed effects, inverse variance weighted methods in the package metafor (v4.6), based on 2-tailed tests. For somatic mutation data, we reported data from up to three measures (presence, burden and activity), as described in the Results. Presence was omitted when the measurement was trivial, *e.g.* all cancers had at least one C:G>A:T mutation). Categorical analyses (mostly 2 x n tables) were generally performed using two-tailed χ^2^ test or, when a value in any cell was <10, by Fisher’s exact test, unless stated to the contrary. For assessing binary explanatory variables and quantitative outcomes, Wilcoxon tests and linear regression were generally used. Linear regression was used for most multivariable analyses. Since many variables were non-normally distributed, models with robust standard errors (using the *lm_robust* command in the *estimatr* package in R) were used for non-normal distributed, proportional or skewed data. Most notably, activities of mutational signatures (although not individual channels such as C:G>T:A) were bimodal, with one maximum at zero and another within an approximately normal distribution at (>0, <1), this being a result of the methods used to derive signatures. ln(odds ratios) (β), standard errors and P values were reported. Covariables in these analyses included combinations of the following: participant age, sex, proximal or distal tumour location (for CRC analyses), hypermutation (from MSI or polymerase mutations), prior cancer genotoxic treatment, TMB, tumour purity, and sequencing from FFPE specimen or non-PCR-free library. We used the R package broom (v1.0.6) to select the combination of variables with the lowest output Akaike information criterion (AIC) value, which best explains the data while limiting overfitting, and combined this with reverse stepwise analysis to ensure that all variables remaining in the final model had nominal P<0.05. In most somatic molecular analyses – such as those involving signature presence, burden and activity – co-variation among both explanatory and outcome variables is both common and complex, and the use of FDR may lead to over-correction. We decided, therefore, to report unadjusted *P* values from 2-tailed tests. As a result, we have endeavoured to interpret association statistics cautiously.

### Declarations

Ethics approval and consent to participate: Ethical approval for collection and analysis of 100kGP patient samples was obtained from the HRA Committee East of England–Cambridge South Research Ethics Committee (REC reference 14/EE/1112). UK Biobank was approved by North West - Haydock Research Ethics Committee 21/NW/0157, IRAS project ID: 299116. The CORGI and CORGI 2 studies were approved by Southampton and SW Hampshire and South Central Research Ethics Committees in the UK with references 06/Q1702/99 and 17/SC/0079. Consent for publication: Not applicable.

### Availability of data and materials

Genomics England permits access to 100kGP data used for this study subject to the following conditions. Research on the de-identified patient data used in this publication can be carried out in the Genomics England Research Environment subject to a collaborative agreement that adheres to patient led governance. All interested readers will be able to access the data in the same manner that the authors accessed the data. For more information about accessing the data, readers may contact or access the relevant information on the Genomics England website, https://www.genomicsengland.co.uk/research. This research has been conducted using the UK Biobank Resource under application numbers 19655 and 155012. UK Biobank data are available through the UK Biobank (http://www.ukbiobank.ac.uk/) upon application, with permission of UKB’s Research Ethics Committee. Other data will be made available by the lead researchers to collaborating researchers, subject to ethical permissions and formal agreement.

## Authors’ contributions

Planned study and wrote manuscript – KS, IT Analysed data – KS, JFT, GG, ST, JW, IT Provided data – JW, CP, TB, RH, MD.

## Data Availability

This research was made possible through access to data in the National Genomic Research Library (https://doi.org/10.6084/m9.figshare.4530893.v7), which is managed by Genomics England Limited (a wholly owned company of the Department of Health and Social Care). The National Genomic Research Library holds data provided by patients and collected by the NHS as part of their care and data collected as part of their participation in research. The National Genomic Research Library is funded by the National Institute for Health Research and NHS England. The Wellcome Trust, Cancer Research UK and the Medical Research Council have also funded research infrastructure. This research has been also been conducted using the UK Biobank Resource under application numbers 19655 and 155012. This manuscript and the underlying study have been made possible partly based on data that Hartwig Medical Foundation has made available to the study through the Hartwig Medical Database under request number DR-219

https://www.genomicsengland.co.uk/

## Acknowledgements

This work made use of the resources provided by the Edinburgh Compute and Data Facility (ECDF) (http://www.ecdf.ed.ac.uk/).

## Supplementary Information

**Supplementary Figure 1.**
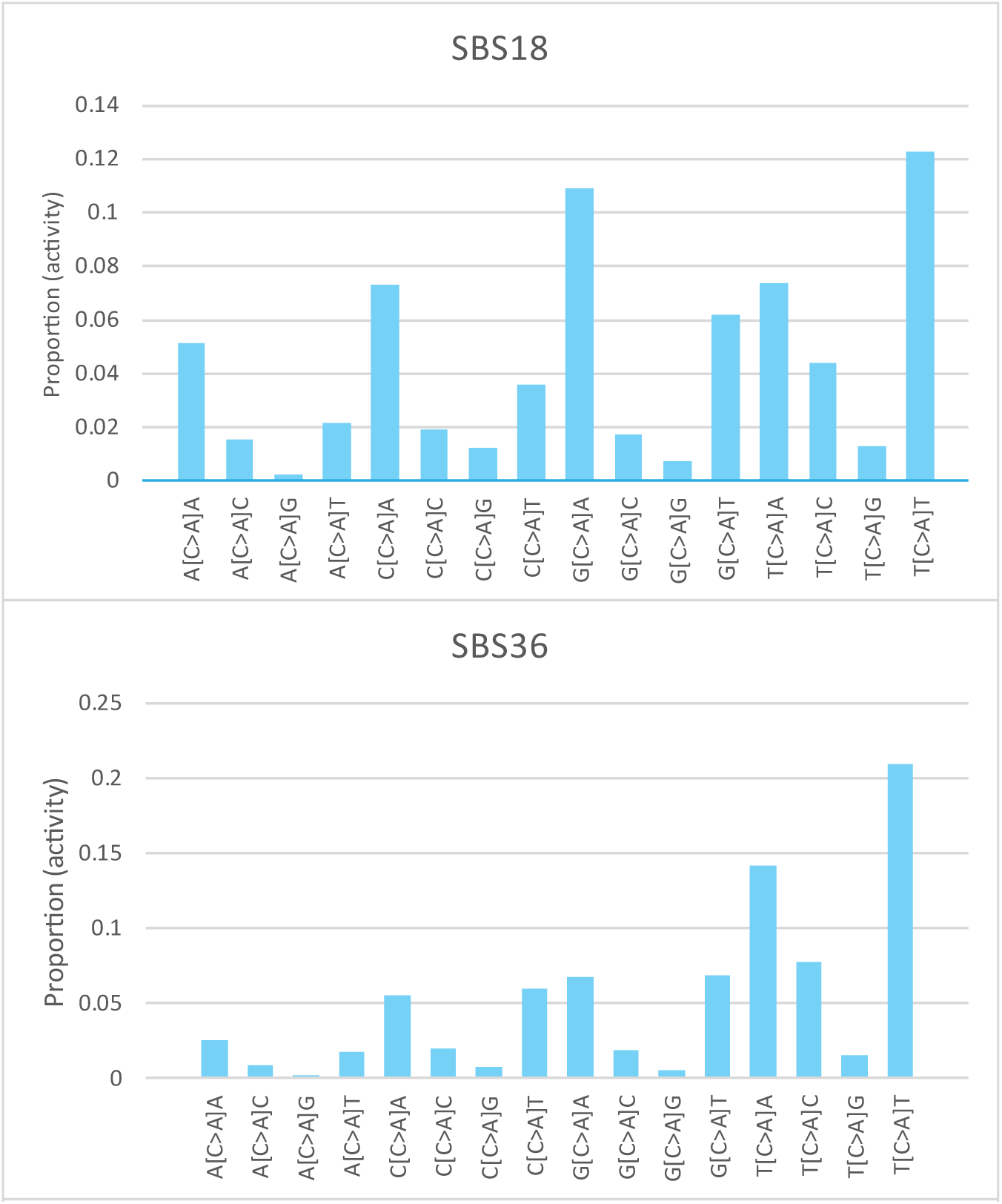
C:G>A>T trinucleotide mutation channels in signatures SBS18 and SBS36.

**Supplementary Figure 2.**
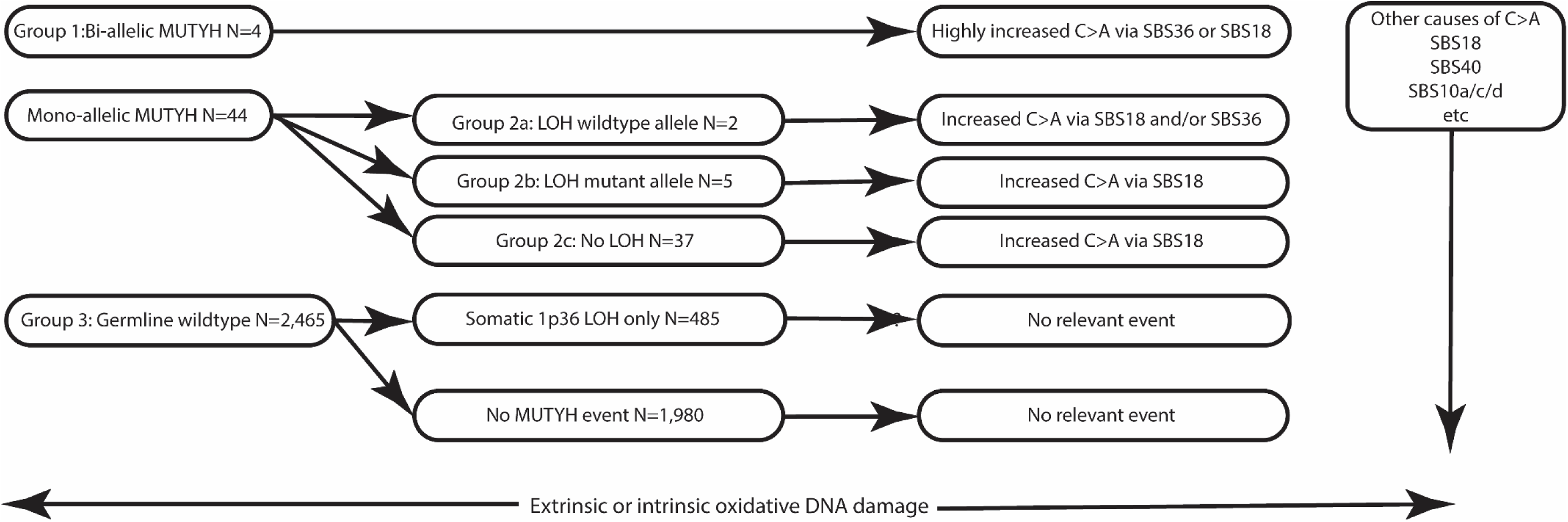
100kGP CRC patients used in somatic molecular analyses, sub-divided by germline MUTYH genotype, second hits at MUTYH and proposed consequences for SBS signatures and C:G>A:T mutations.

**Supplementary Figure 3.**
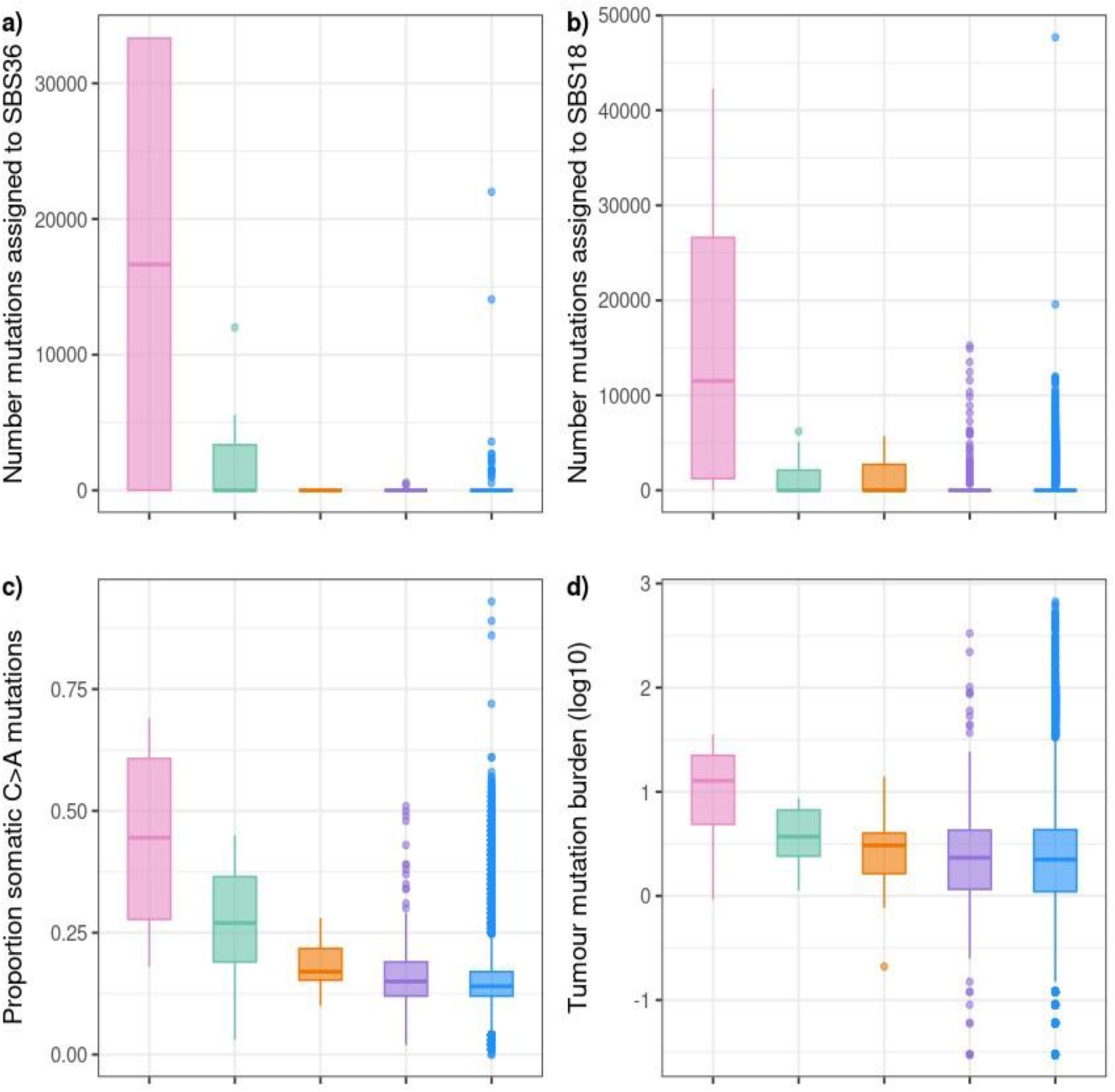
Selected somatic molecular features of tumours in the 100kGP All-cancer analysis in relation to germline MUTYH status. (a) SBS36 proportional activity, (b) SBS18 proportion activity, (c) C:G>A:T burden, (d) C:G>A:T proportional activity and (e) tumour mutational burden (measured as SCVPM) in germline MUTYH genotype groups 1, 2a, 2b, 2c and 3. Group 1: bi-allelic germline MUTYH mutations; group 2a: mono-allelic germline MUTYH mutation with LOH wildtype allele; group 2b: mono-allelic germline *MUTYH* mutation with LOH mutant allele; group 2c: mono-allelic germline MUTYH mutation with no LOH wildtype; group 3: germline MUTYH wildtype. Log scales are used to aid display in some cases. Data and association statistics corresponding to the above are shown in **Table 1** and **Supplementary Tables 1-7.**

**Supplementary Table 1.** Summary of somatic molecular features of 100kGP CRCs. μ mean, σ standard deviation, M median. SCVPM, somatic coding variants per Mb, a measure of tumour mutation burden focussed on variants more likely to have pathogenic effects. Group 1: bi-allelic germline *MUTYH* mutations; group 2a: mono-allelic germline *MUTYH* mutations with LOH of germline wildtype allele; group 2b: mono-allelic germline *MUTYH* mutations with LOH of germline mutant allele; group 2c: mono-allelic germline *MUTYH* mutations with no LOH; group 3: wildtype *MUTYH* alleles.

| Molecular feature |  | Group 1 | Group 2a | Group 2b | Group 2c | Group 3 |
| --- | --- | --- | --- | --- | --- | --- |
|  | n | 4 | 2 | 5 | 37 | 2465 |
| Age | $\mu$ | 52 | 71 | 68 | 67 | 68 |
| Presence SBS36 | $\mu$ | 0.75 | 0.50 | 0.00 | 0.00 | 0.00 |
| | $\sigma$ | 0.50 | 0.71 | 0.00 | 0.00 | 0.06 |
|  | M | 1.00 | 0.50 | 0.00 | 0.00 | 0.00 |
| Burden SBS36 | $\mu$ | 24366 | 5999 | 0 | 0 | 17 |
| | $\sigma$ | 16290 | 8484 | 0 | 0 | 464 |
|  | M | 32033 | 5999 | 0 | 0 | 0 |
| Activity SBS36 | $\mu$ | 0.377 | 0.231 | 0.000 | 0.000 | 0.001 |
| | $\sigma$ | 0.372 | 0.327 | 0.000 | 0.000 | 0.013 |
|  | M | 0.327 | 0.231 | 0.000 | 0.000 | 0.000 |
| Presence SBS18 | $\mu$ | 0.50 | 0.50 | 0.80 | 0.46 | 0.27 |
| | $\sigma$ | 0.58 | 0.71 | 0.45 | 0.51 | 0.44 |
|  | M | 0.50 | 0.50 | 1.00 | 0.00 | 0.00 |
| Burden SBS18 | $\mu$ | 15914 | 2117 | 3461 | 2689 | 1146 |
| | $\sigma$ | 20258 | 2994 | 2190 | 3610 | 2307 |
|  | M | 10692 | 2117 | 3833 | 0 | 0 |
| Activity SBS18 | $\mu$ | 0.171 | 0.131 | 0.176 | 0.115 | 0.054 |
| | $\sigma$ | 0.201 | 0.195 | 0.101 | 0.134 | 0.093 |
|  | M | 0.151 | 0.131 | 0.214 | 0.000 | 0.000 |
| Burden C>A | $\mu$ | 34464 | 8249 | 4035 | 6739 | 9542 |
| | $\sigma$ | 6067 | 4846 | 1140 | 7330 | 44828 |
|  | M | 34765 | 8249 | 4186 | 3980 | 3668 |
| Activity C>A | $\mu$ | 0.448 | 0.379 | 0.231 | 0.203 | 0.173 |
| | $\sigma$ | 0.223 | 0.101 | 0.030 | 0.070 | 0.050 |
|  | M | 0.448 | 0.379 | 0.218 | 0.193 | 0.172 |
| SCVPM (TMB) | $\mu$ | 36.4 | 5.2 | 3.4 | 12.33 | 17.49 |
| | $\sigma$ | 32.7 | 1.9 | 1.6 | 25.65 | 36.33 |
|  | M | 27.3 | 5.2 | 4.1 | 3.80 | 3.86 |

**Supplementary Table 2.**
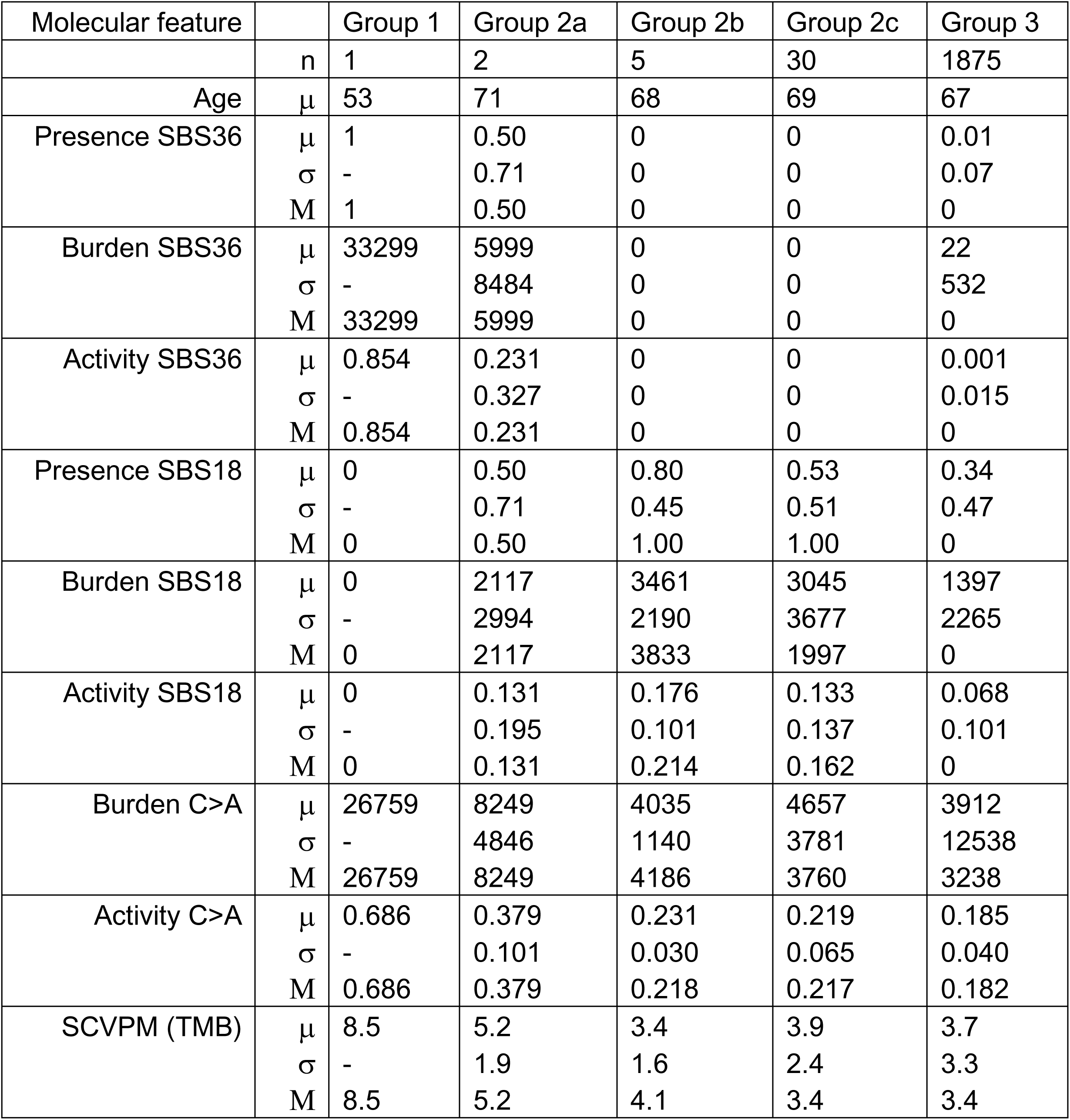
Summary of somatic molecular features of 100kGP MSS CRCs. Legend as in **Supplementary Table 2**.

**Supplementary Table 3.** SBS36 & SBS18 presence & absence in germline MUTYH mutation groups in cases with CRC, ‘All-Cancer’, and ‘CRC-excluded’ from the 100kGP. The upper table shows tumour counts; the lower table shows associations with *MUTYH* groups (1=bi-allelic germline mutations, 2a=mono-allelic germline mutation + loss of the wildtype allele, 2b= mono-allelic germline mutation + loss of the mutant allele, 2c= mono-allelic germline mutation with no loss, 3=germline MUTYH-wildtype). ND=not determined.

| Tumour type | Germline *MUTYH* group | SBS36 present | SBS36 absent | Total | SBS18 present | SBS18 absent | Total |
| --- | --- | --- | --- | --- | --- | --- | --- |
| CRC | 1 | 3 | 1 | 4 | 2 | 2 | 4 |
|  | 2a | 1 | 1 | 2 | 1 | 1 | 2 |
|  | 2b | 0 | 5 | 5 | 4 | 1 | 5 |
|  | 2c | 0 | 37 | 37 | 17 | 20 | 37 |
|  | All mono-allelic | 1 | 43 | 44 | 22 | 22 | 44 |
|  | 3 | 10 | 2455 | 2465 | 669 | 1796 | 2465 |
|  | Total | 14 | 2499 | 2513 | 693 | 1820 | 2513 |
| All-cancer | 1 | 3 | 2 | 5 | 3 | 2 | 5 |
|  | 2a | 5 | 7 | 12 | 3 | 9 | 12 |
|  | 2b | 0 | 14 | 14 | 5 | 9 | 14 |
|  | 2c | 3 | 242 | 245 | 46 | 199 | 245 |
|  | All mono-allelic | 8 | 263 | 271 | 57 | 219 | 271 |
|  | 3 | 18 | 14929 | 14947 | 1232 | 13715 | 14947 |
|  | Total | 29 | 15194 | 15223 | 1289 | 13934 | 15223 |
| CRC excluded | 1 | 0 | 1 | 1 | 1 | 0 | 1 |
|  | 2a | 4 | 6 | 10 | 2 | 8 | 10 |
|  | 2b | 0 | 9 | 9 | 1 | 9 | 9 |
|  | 2c | 3 | 199 | 202 | 26 | 176 | 202 |
|  | All mono-allelic | 7 | 214 | 221 | 29 | 192 | 221 |
|  | 3 | 8 | 12304 | 12312 | 530 | 11782 | 12312 |
|  | Total | 15 | 12519 | 12534 | 560 | 11974 | 12534 |

|  | Signature | MUTYH groups | Odds ratio | 95% CI | P | Test |
| --- | --- | --- | --- | --- | --- | --- |
| CRC | SBS36 | 1 vs 2a/b/c+3 | 681 | 65.7-7067 | 1.10x10<sup>-6</sup> | Fisher's exact |
|  |  | 2a vs 2b/c | ND | ND | 0.090 | Fisher's exact |
|  |  | 2a/b/c vs 3 | 5.71 | 0.72-46 | 0.354 | χ<sup>2</sup> |
|  | SBS18 | 1 vs 2a/b/c+3 | 2.63 | 0.37-19 | 0.612 | Fisher's exact |
|  |  | 2a vs 2b/c | 1 | 0.06-17 | 1 | Fisher's exact |
|  |  | 2a/b/c vs 3 | 2.685 | 1.51-4.76 | 4.48x10<sup>-4</sup> | χ<sup>2</sup> |
| All-cancer | SBS36 | 1 vs 2a/b/c+3 | 141 | 141-5464 | 1.24x10<sup>-7</sup> | Fisher's exact |
|  |  | 2a vs 2b/c | 61 | 12.1-307 | 7.07x10<sup>-6</sup> | Fisher's exact |
|  |  | 2a/b/c vs 3 | 25.2 | 10.9-59 | <10<sup>-7</sup> | χ<sup>2</sup> |
|  | SBS18 | 1 vs 2a/b/c+3 | 16.7 | 2.79-100 | 9.90x10<sup>-4</sup> | Fisher's exact |
|  |  | 2a vs 2b/c | 1.36 | 3.55-320 | 0.881 | Fisher's exact |
|  |  | 2a/b/c vs 3 | 2.90 | 2.15-3.90 | <10<sup>-7</sup> | χ<sup>2</sup> |
| CRC excluded | SBS36 | 1 vs 2a/b/c+3 | 1 | ND | 1 | Fisher's exact |
|  |  | 2a vs 2b/c | 46 | 8.42-254 | 1.42x10<sup>-4</sup> | Fisher's exact |
|  |  | 2a/b/c vs 3 | 50 | 18.1-140 | <10<sup>-7</sup> | Fisher's exact |
|  | SBS18 | 1 vs 2a/b/c+3 | 1 | ND | 0.089 | Fisher's exact |
|  |  | 2a vs 2b/c | 1.71 | 0.345-8.49 | 0.769 | Fisher's exact |
|  |  | 2a/b/c vs 3 | 3.36 | 1.76-5.52 | 3.44x10<sup>-7</sup> | χ<sup>2</sup> |

**Supplementary Table 4.** Pairwise associations in univariable analyses between germline MUTYH genotypes and cancer molecular features in 100kGP CRCs, All-cancer, and CRC-excluded tumour sets. For binary variables, ORs & 95% CIs are from 2x2 tables & P values from χ^2^ or Fisher’s exact tests, whereas for associations involving quantitative variables (burden & activity), P values are from Wilcoxon tests. Effect size metrics (i.e. βs (log(ORs)), standard errors (SE) & corresponding P_regress_ values) are estimated from univariable regression with robust variances. ND, tests not performed owing to small sample numbers.

| Groups | Somatic features | CRC only |  |  |  | All-cancer |  |  |  | CRC-excluded |  |  |  |
| --- | --- | --- | --- | --- | --- | --- | --- | --- | --- | --- | --- | --- | --- |
| | | P | $\beta$ | SE | $P_{\text{regress}}$ | P | $\beta$ | SE | $P_{\text{regress}}$ | P | $\beta$ | SE | $P_{\text{regress}}$ |
| 1 vs 2+3 | Presence SBS36 | 1.10x10<sup>-6</sup> | 0.746 | 0.034 | <2.2x10<sup>-16</sup> | 1.24x10<sup>-7</sup> | 0.598 | 0.019 | <2.2x10<sup>-16</sup> | 1 | ND | ND | ND |
|  | Burden SBS36 | 0.00080 | 24345 | 8145 | 0.00282 | <2.2x10<sup>-16</sup> | 19487 | 7972 | 0.015 | ND | ND | ND | ND |
|  | Activity SBS36 | 0.0016 | 0.376 | 0.186 | 0.0434 | <2.2x10<sup>-16</sup> | 0.301 | 0.163 | 0.064 | ND | ND | ND | ND |
|  | Presence SBS18 | 0.61 | 0.225 | 0.224 | 0.315 | 0.011 | 0.515 | 0.124 | 3.46x10<sup>-5</sup> | 0.089 | ND | ND | ND |
|  | Burden SBS18 | 0.11 | 14740 | 10129 | 0.150 | 1.47x10<sup>-5</sup> | 12764 | 8349 | 0.126 | ND | ND | ND | ND |
|  | Activity SBS18 | 0.12 | 0.116 | 0.100 | 0.250 | 1.24x10<sup>-5</sup> | 0.155 | 0.078 | 0.047 | ND | ND | ND | ND |
|  | Burden C>A | 0.00080 | 24975 | 3161 | 4.08x10<sup>-15</sup> | 0.0026 | 21476 | 6958 | 2.03x10<sup>-3</sup> | ND | ND | ND | ND |
|  | Activity C>A | 0.0016 | 0.275 | 0.112 | 0.014 | 0.0011 | 0.237 | 0.102 | 0.020 | ND | ND | ND | ND |
|  | TMB | <2.2x10<sup>-16</sup> | 19.0 | 16.37 | 0.245 | 0.025 | 21.3 | 14.5 | 0.143 | ND | ND | ND | ND |
| 2a vs 2b/c | Presence SBS36 | 0.091 | ND | ND | ND | 7.07x10<sup>-6</sup> | 0.405 | 0.044 | <2.2x10<sup>-16</sup> | 1.42x10<sup>-4</sup> | 0.386 | 0.164 | 0.019 |
|  | Burden SBS36 | 7.70x10<sup>-6</sup> | ND | ND | ND | 2.87x10<sup>-16</sup> | 2134 | 1052 | 0.043 | 3.30x10<sup>-12</sup> | 1361 | 653 | 0.038 |
|  | Activity SBS36 | 7.70x10<sup>-6</sup> | ND | ND | ND | 2.87x10<sup>-16</sup> | 0.121 | 0.053 | 0.024 | 3.30x10<sup>-12</sup> | 0.099 | 0.052 | 0.057 |
|  | Presence SBS18 | 1.00 | ND | ND | ND | 0.881 | 0.053 | 0.118 | 0.654 | 0.77 | 0.072 | 0.135 | 0.594 |
|  | Burden SBS18 | 0.90 | ND | ND | ND | 0.569 | 289 | 710 | 0.684 | 0.51 | 593 | 772 | 0.443 |
|  | Activity SBS18 | 0.81 | ND | ND | ND | 0.558 | 0.020 | 0.034 | 0.569 | 0.52 | 0.024 | 0.034 | 0.473 |
|  | Burden C>A | 0.25 | ND | ND | ND | 0.031 | -2499 | 3536 | 0.480 | 0.040 | -3335 | 4264 | 0.435 |
|  | Activity C>A | 0.026 | ND | ND | ND | 0.0062 | 0.086 | 0.039 | 0.026 | 0.023 | 0.071 | 0.041 | 0.085 |
|  | TMB | 0.38 | ND | ND | ND | 0.141 | -3.48 | 1.87 | 0.064 | 0.185 | -2.67 | 2.11 | 0.207 |
| 2 vs 3 | Presence SBS36 | 0.35 | 0.187 | 0.010 | 0.063 | <10<sup>-7</sup> | 0.028 | 0.003 | <2.2x10<sup>-16</sup> | <10<sup>-7</sup> | 0.031 | 0.012 | 0.009 |
|  | Burden SBS36 | 0.062 | 255.9 | 273 | 0.348 | <2.2x10<sup>-16</sup> | 95.6 | 52.2 | 0.067 | <2.2x10<sup>-16</sup> | 66.4 | 34.2 | 0.052 |
|  | Activity SBS36 | 0.062 | 0.0099 | 0.011 | 0.348 | <2.2x10<sup>-16</sup> | 0.006 | 0.003 | 0.028 | <2.2x10<sup>-16</sup> | 0.005 | 0.003 | 0.041 |
|  | Presence SBS18 | 4.48x10<sup>-4</sup> | 0.229 | 0.068 | 7.61x10<sup>-4</sup> | <10<sup>-7</sup> | 0.117 | 0.017 | 6.98x10<sup>-12</sup> | 4.44x10<sup>-7</sup> | 0.088 | 0.023 | 1.13x10<sup>-4</sup> |
|  | Burden SBS18 | 9.59x10<sup>-5</sup> | 1605 | 517 | 1.99x10<sup>-3</sup> | 2.19x10<sup>-12</sup> | 735 | 176 | 2.98x10<sup>-5</sup> | 8.88x10<sup>-12</sup> | 462 | 143 | 1.26x10<sup>-3</sup> |
|  | Activity SBS18 | 2.61x10<sup>-5</sup> | 0.069 | 0.020 | 5.70x10<sup>-4</sup> | 1.18x10<sup>-12</sup> | 0.029 | 0.006 | 1.05x10<sup>-6</sup> | 8.60x10<sup>-12</sup> | 0.018 | 0.005 | 1.16x10<sup>-4</sup> |
|  | Burden C>A | 0.58 | -3041 | 1369 | 0.026 | 0.045 | 833 | 3207 | 0.795 | 0.016 | 1648 | 3922 | 0.674 |
|  | Activity C>A | 5.19x10<sup>-5</sup> | 0.041 | 0.012 | 3.65x10<sup>-4</sup> | 1.22x10<sup>-6</sup> | 0.0180 | 0.005 | 3.78x10<sup>-4</sup> | 2.02x10<sup>-5</sup> | 0.012 | 0.005 | 0.027 |
|  | TMB | 0.34 | -6.50 | 3.64 | 0.075 | 0.811 | -0.732 | 1.652 | 0.658 | 0.445 | 0.334 | 1.851 | 0.857 |

**Supplementary Table 5.** Multivariable regression analysis for associations between germline MUTYH genotypes and mutational processes in 100kGP CRCs. Generalised linear models (glm) were used for binary outcome variables and linear regression with robust variances for quantitative outcomes. Included as co-variables in each initial regression model were tumour location (proximal=between caecum and splenic flexure, distal=between descending colon and rectum), MSI, primary or metastasis, age, sex, tumour purity, prior chemotherapy, prior radiotherapy and TMB defined as somatic coding variants per Mb (unless an outcome). Non-significant variables at P=0.05 were sequentially excluded in reverse stepwise regression analysis until all variables with P<0.05 remained. Where the *MUTYH* group variable remained significantly associated with molecular features (P<0.05), other positively associated co-variables in the final model are shown. Where the *MUTYH* variable was excluded from the final model (P>0.05), NS is shown. The table complements **Table 2** and **Supplementary Tables 4 & 5**. **Supplementary Tables 17 & 19** provide equivalent data for All-cancer and CRC-excluded analyses.

| Groups | Somatic features | $\beta$ | SE | P | Positively associated co-variables, $P<0.05$ |
| --- | --- | --- | --- | --- | --- |
| 1 v 2+3 | Presence SBS36 | 0.749 | 0.249 | 0.0027 | MSS |
|  | Burden SBS36 | 24345 | 8145 | 0.0028 |  |
|  | Activity SBS36 | 0.377 | 0.186 | 0.043 | MSS |
|  | Presence SBS18 |  |  | NS |  |
|  | Burden SBS18 |  |  | NS |  |
|  | Activity SBS18 |  |  | NS |  |
|  | Burden C>A |  |  | NS |  |
|  | Activity C>A | 0.280 | 0.107 | 0.0090 | Low TMB |
|  | TMB |  |  | NS |  |
| 2 v 3 | Presence SBS36 |  |  | NS |  |
|  | Burden SBS36 |  |  | NS |  |
|  | Activity SBS36 |  |  | NS |  |
| | Presence SBS18 | 0.244 | 0.075 | $9.27 \times 10^{-4}$ | MSS, proximal location, primary, sex |
| | Burden SBS18 | 1741 | 511 | $6.80 \times 10^{-4}$ | MSS, proximal location, primary |
| | Activity SBS18 | 0.081 | 0.022 | $2.22 \times 10^{-4}$ | MSS, proximal location, primary |
|  | Burden C>A | 1639 | 833 | 0.049 | MSS, proximal location |
| | Activity C>A | 0.042 | 0.010 | $5.69 \times 10^{-5}$ | MSS, proximal location, age |
|  | TMB |  |  | NS |  |

**Supplementary Table 6.** Univariable associations between MUTYH genotype (group 2 vs group 3) and (a) CRC location (proximal v distal colorectum) and (b) MSI. Despite their presence as significant predictors in the multivariable models, proximal location and MSS status were not associated with group 2 in the univariable analyses *versus* group3.

|  |  | Proximal | Distal | Total |  | MSI | MSS | Total |
| --- | --- | --- | --- | --- | --- | --- | --- | --- |
| *MUTYH* mono-allelic (group 2) | a | 12 | 30 | 42 | b | 7 | 37 | 44 |
| Wildtype (group 3) |  | 979 | 1447 | 2426 |  | 569 | 1875 | 2444 |
| Total |  | 991 | 1477 | 2468 |  | 576 | 1912 | 2488 |

(a) OR=0.591, 95%CI 0.301-1.16; $P=0.162$ , Fisher's exact test
(b) OR=0.623, 95%CI 0.276-1.406; $P=0.332$ , Fisher's exact test

**Supplementary Table 7.**
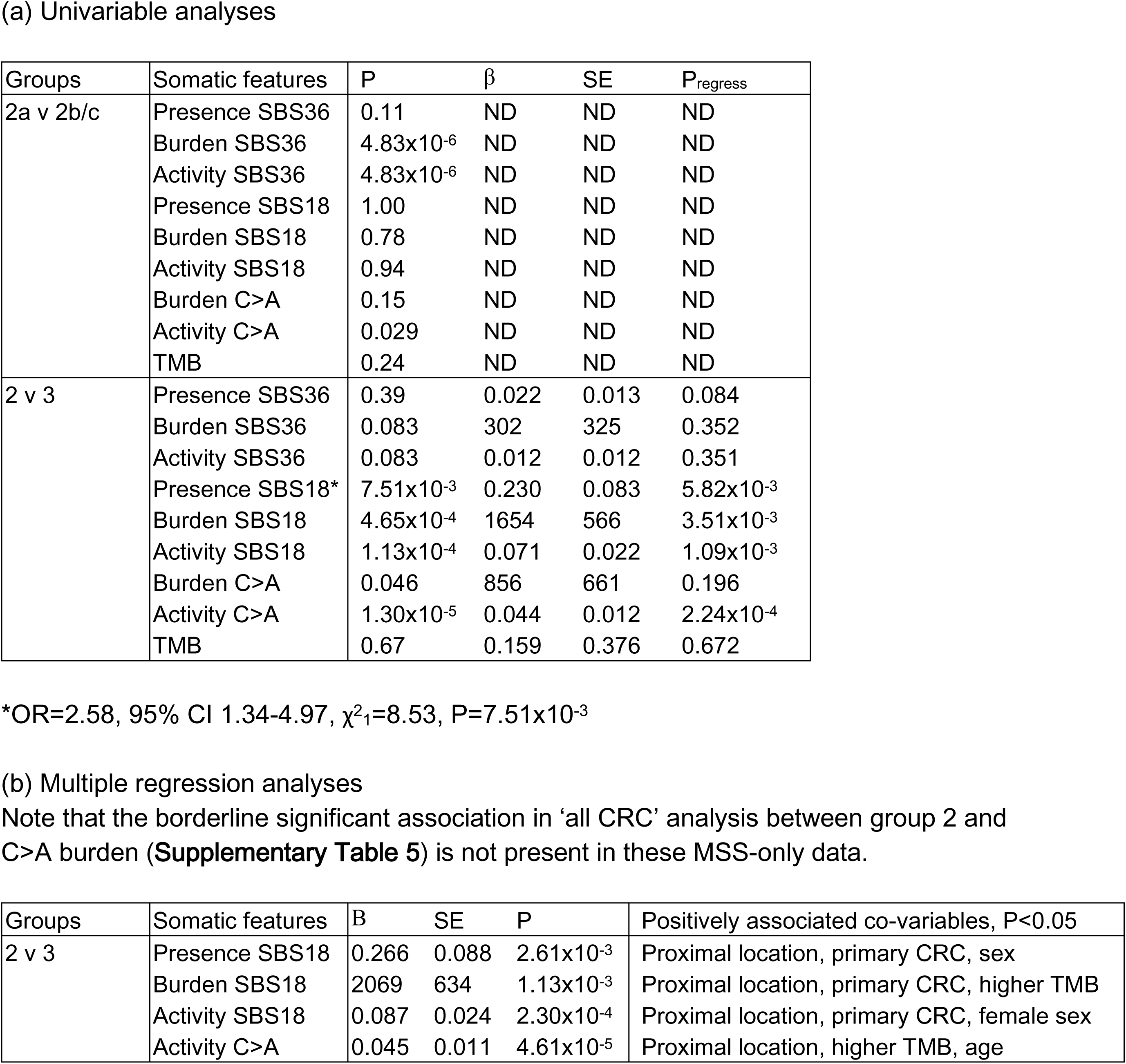
Associations between germline MUTYH genotypes and molecular features in 100kGP MSS CRC-only analyses. MSI+ and POL-mutant cancers were excluded. As per **Supplementary Table 5,** for binary variables, ORs and 95% CIs are from 2x2 tables and P values from χ^2^ or Fisher’s exact tests, whereas for associations involving quantitative variables (burden and activity), P values are from Wilcoxon tests. Effect size metrics (i.e. βs (log(ORs)), standard errors (SE) and corresponding P_regress_ values) are estimated from univariable regression with robust variances. ND shows tests not performed owing to small sample numbers. Other annotation is as per **Supplementary Tables 4-6**. Group 2a: mono-allelic germline *MUTYH* mutation with LOH wildtype allele; group 2b: mono-allelic germline *MUTYH* mutation with LOH mutant allele; group 2c: mono-allelic germline *MUTYH* mutation with no LOH wildtype; group 3: germline *MUTYH* wildtype

**Supplementary Table 8.** Comparison between SBS18-positive CRCs from mono-allelic MUTYH mutation carriers (n=17, group 2) and MUTYH-wildtype individuals (n=528, group 3). Median values are shown for quantitative variables, and numbers of tumours/total for binary variables (location, whole genome doubling, MSI, primary or metastasis). P values are derived from Wilcoxon tests or Fisher’s exact tests. This analysis was performed on a sub-set of individuals from whom data such as whole genome doubling were available.

|  | *Group 2* | *Group 3* | *P* |
| --- | --- | --- | --- |
| SBS18 burden | 5729 | 3859 | 0.0029 |
| SBS18 activity | 0.250 | 0.197 | 0.00019 |
| C>A burden | 5326 | 3904 | 0.0014 |
| C>A activity | 0.247 | 0.197 | 2.37x10<sup>-4</sup> |
| SCVPM | 4.20 | 3.71 | 0.120 |
| Age (years) | 71 | 70 | 0.803 |
| Location (proximal colorectum) | 11/17 | 263/528 | 0.336 |
| Whole genome doubling | 7/16 | 201/467 | 0.129 |
| MSI | 0/17 | 2/526 | 1.000 |
| Primary | 17/17 | 503/528 | 0.889 |

**Supplementary Table 9.**
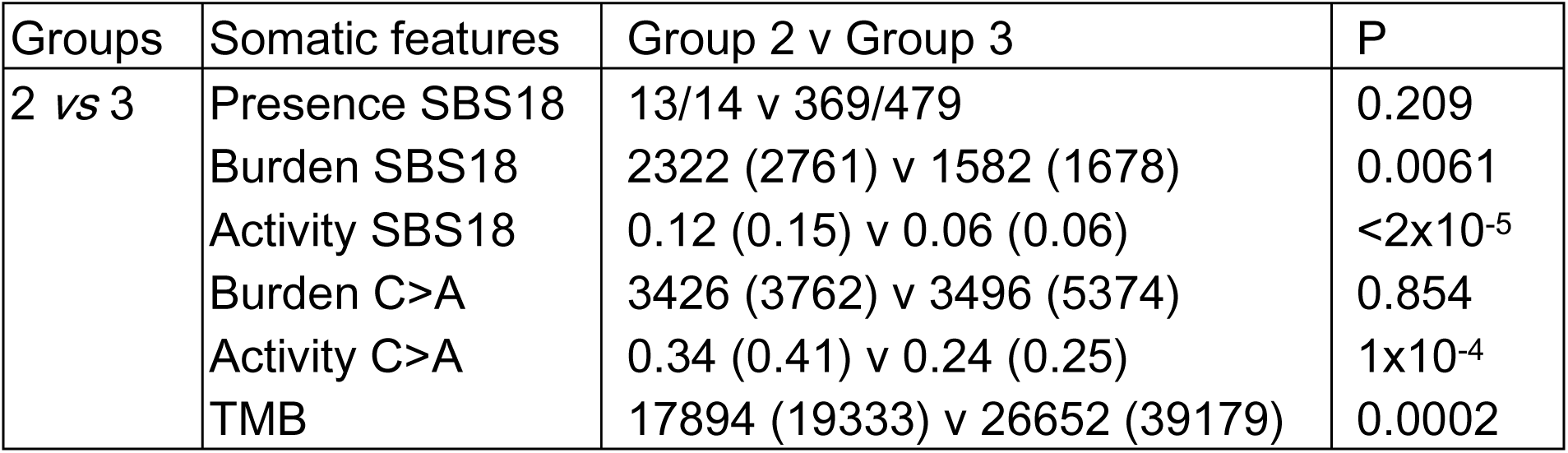
Associations between mono-allelic germline MUTYH mutations (group 2) and molecular features in 493 metastatic CRCs from Hartwig study. Fourteen CRCs were from germline mono-allelic mutation carriers. Data are from Fisher’s exact test (for presence/absence) and from WIlcoxon tests (for all other measures). Median values are shown for burdens and activities, with means in brackets. Here, owing to the data available, TMB was expressed as the sum of all SBS mutations. MSS and MSI data were combined for this analysis, with all 23 MSI+ cancers in the *MUTYH-*wildtype group (hence the lower between TMB in group 2 cancers), and many cancers were sampled after receiving chemotherapy, which is likely to have introduced background mutational ‘noise’. For these reasons, as for the 100kGP analyses, we regarded SBS18 as the most reliable measure of MUTYH function here, whereas measures of mutation burden were subject to additional factors of uncertain magnitude. We were reassured that most SBS18 measures, including burden, were associated with mono-allelic *MUTYH* carriers, as was C>A activity.

**Supplementary Table 10.** Exome SNP array data for MUTYH p.Gly382Asp and Tyr179Cys. A total of 20 *MUTYH* variants, all of which were missense or possible splice changes and mostly predicted to be non-pathogenic, was present on these SNP arrays. p.Tyr179Cys and p.Gly396Asp were the only pathogenic variants found in our patients. Whilst a small number of homozygous p.Tyr179Cys and p.Gly396Asp carriers were found, there was no compound heterozygote p.Tyr179Cys/p.Gly396Asp individual. We cannot exclude, for example, that a small number (probably <1%) of the apparent mono-allelic p.Tyr179Cys or p.Gly396Asp mono-allelic mutants were actually compound heterozygotes with a second, rare, untyped pathogenic variant, and hence these data do not form part of the formal meta-analyses in **Table 1**.

|  | N mono-allelic cases/total | N mono-allelic controls/total | OR | 95%CI | P |
| --- | --- | --- | --- | --- | --- |
| G396D/+ | 89/5554 | 90/8091 | 1.448 | 1.078-1.945 | 0.0174 |
| Y165C/+ | 7/5574 | 14/8069 | 0.7235 | 0.292-1.793 | 0.640 |

**Supplementary Table 11.** Exploration of potential confounders that could have caused the association between mono-allelic MUTYH mutations and SBS18 burden in 100kGP CRCs. Results are from a multivariable linear model including only MSS, POL-proficient CRCs. SBS18 burden was the outcome and *MUTYH* groups 2 *versus* 3 (germline heterozygotes *versus* wildtype) were the main explanatory variable, with each co-variable included in turn. All models also included sex and age. Whilst some variables were significantly associated with SBS18 burden, the impact of the mono-allelic *MUTYH* genotype was still significant in all instances. Note that the association with ancestry was driven by a single individual of African descent. BER genes tested were *OGG1, UNG, SMUG1, TDG, NTHL1, MPG, NEIL1, NEIL2, NEIL3, PARP1, PARP2, PARP3, PARG* and *PARPBP.* Associations with age, sex and location were detected previously in other multivariable analyses (see above). Note that although not shown in the table, there was no association between SBS18 and measures of the microbiome, as previously reported by Cornish *et al* ^27^.

| Co-variable | $\beta$ | SE | P | Significance of *MUTYH* genotype after controlling for co-variable |
| --- | --- | --- | --- | --- |
| Participant age | 20.049 | 4.591 | 1.33x10<sup>-5</sup> | 1.86x10<sup>-5</sup> |
| Participant sex | -259.060 | 108.71 | 0.017 | 1.61x10<sup>-5</sup> |
| Prior chemotherapy or radiotherapy | -123.192 | 109.155 | 0.259 | 2.00x10<sup>-5</sup> |
| Proximal vs distal colorectum location | 1532.084 | 107.961 | <2e<sup>-16</sup> | 4.15x10<sup>-7</sup> |
| Tumour type - primary vs met | 509.171 | 212.68 | 0.017 | 2.08x10<sup>-5</sup> |
| Tumour stage (1-4) | -16.804 | 63.356 | 0.791 | 8.53x10<sup>-6</sup> |
| Genetic ancestry – African vs European | 1272.187 | 351.148 | 2.99x10<sup>-4</sup> | 1.91x10<sup>-5</sup> |
| Genetic ancestry – American vs European | -884.907 | 328.171 | 0.007 | 1.91x10<sup>-5</sup> |
| Genetic ancestry – East Asian vs European | -608.782 | 571.328 | 0.287 | 1.91x10<sup>-5</sup> |
| Genetic ancestry – Mixed vs European | -13.181 | 165.507 | 0.937 | 1.91x10<sup>-5</sup> |
| Tumour purity (ASCAT) | -10.157 | 238.683 | 0.970 | 2.06x10<sup>-5</sup> |
| Tumour ploidy (ASCAT) | -59.761 | 63.045 | 0.343 | 2.73x10<sup>-5</sup> |
| Presence of other BER mutation | 87.909 | 468.086 | 0.851 | 2.15x10<sup>-5</sup> |

**Supplementary Table 12.**
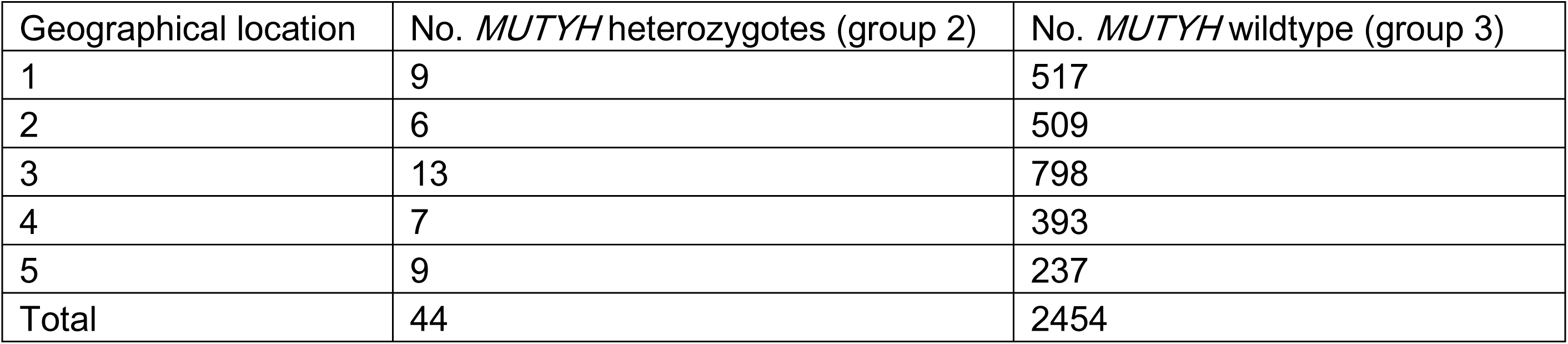
Geographical origins of the patients from different regions of England. 100kGP participants were recruited by one of 12 Genomic Medicine Centres. In order to create large enough sets of patients of some types, we reduced the number of locations to five by merging some geographically adjacent centres (e.g. London centres were combined). Centre identities are not provided for reasons of confidentiality. There was no association between *MUTYH* heterozygosity and location of the patient recruitment centre (heterogeneity χ^2^, P=0.178).

**Supplementary Table 13.**
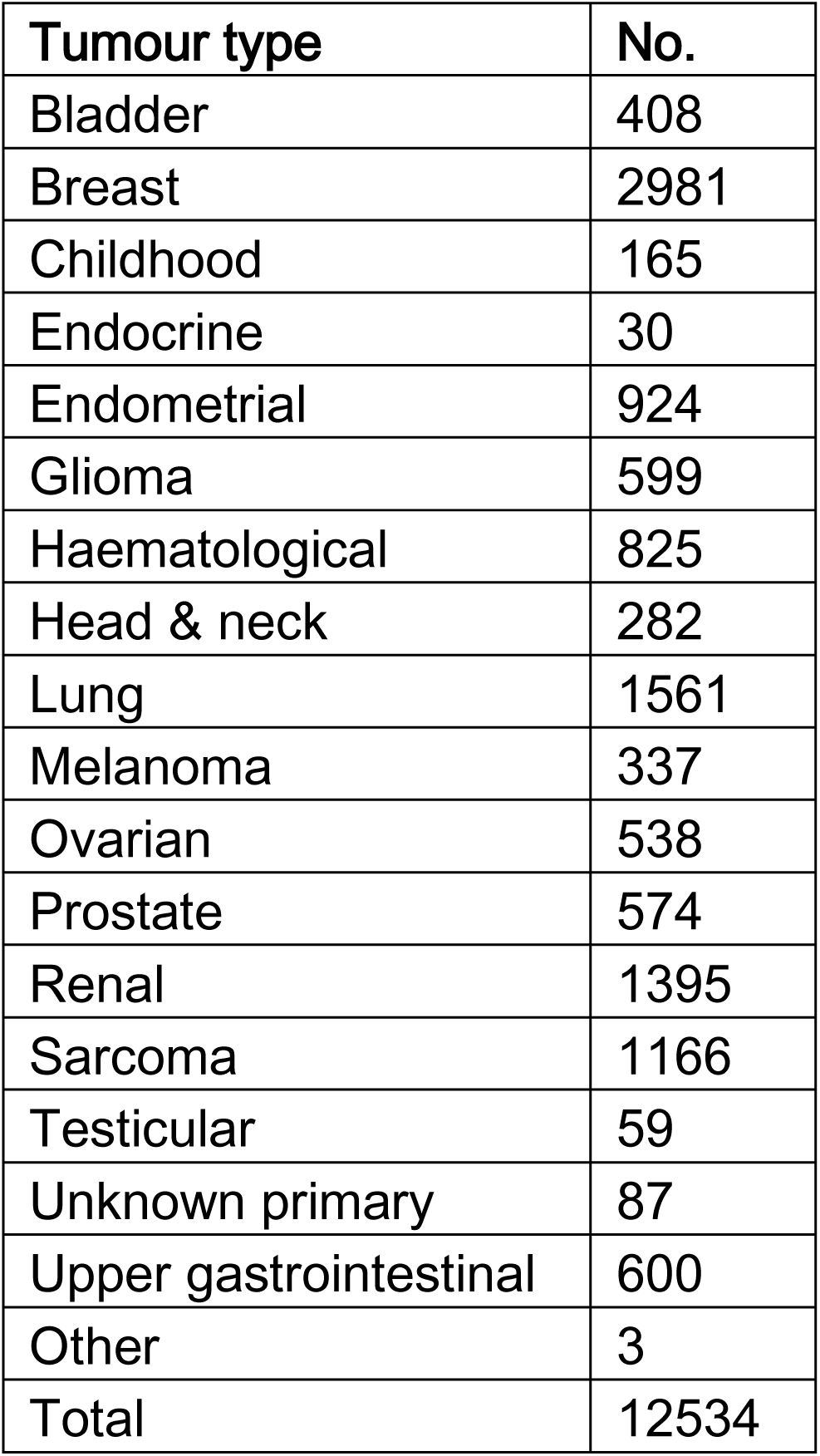
Non-CRC cancers analysed for associations between MUTYH genotypes and somatic molecular features.

**Supplementary Table 14.** Summary of somatic molecular features of 100kGP All-cancer tumours. Legend as in **Supplementary Table 3**.

| Molecular feature |  | Group 1 | Group 2a | Group 2b | Group 2c | Group 3 |
| --- | --- | --- | --- | --- | --- | --- |
|  | n | 5 | 12 | 14 | 245 | 14947 |
| Age | $\mu$ | 48 | 55 | 68 | 62 | 62 |
| Presence SBS36 | $\mu$ | 0.60 | 0.42 | 0 | 0.01 | 0 |
| | $\sigma$ | 0.55 | 0.51 | 0 | 0.11 | 0.03 |
|  | M | 1.00 | 0 | 0 | 0 | 0 |
| Burden SBS36 | $\mu$ | 19493 | 2140 | 0 | 6 | 4 |
| | $\sigma$ | 17826 | 3644 | 0 | 52 | 223 |
|  | M | 30766 | 0 | 0 | 0 | 0 |
| Activity SBS36 | $\mu$ | 0.301 | 0.122 | 0 | 0.001 | 0 |
| | $\sigma$ | 0.364 | 0.184 | 0 | 0.009 | 0.007 |
|  | M | 0.184 | 0 | 0 | 0 | 0 |
| Presence SBS18 | $\mu$ | 0.60 | 0.25 | 0.36 | 0.19 | 0.08 |
| | $\sigma$ | 0.55 | 0.45 | 0.50 | 0.39 | 0.28 |
|  | M | 1.00 | 0 | 0 | 0 | 0 |
| Burden SBS18 | $\mu$ | 13060 | 1295 | 1372 | 984 | 283 |
| | $\sigma$ | 18670 | 2379 | 2082 | 2961 | 1184 |
|  | M | 1641 | 0 | 0 | 0 | 0 |
| Activity SBS18 | $\mu$ | 0.170 | 0.063 | 0.072 | 0.041 | 0.015 |
| | $\sigma$ | 0.174 | 0.117 | 0.104 | 0.096 | 0.054 |
|  | M | 0.167 | 0 | 0 | 0 | 0 |
| Burden C>A | $\mu$ | 27922 | 4875 | 3104 | 7618 | 6431 |
| | $\sigma$ | 15542 | 4037 | 2335 | 55234 | 38834 |
|  | M | 34294 | 4240 | 2408 | 1864 | 1671 |
| Activity C>A | $\mu$ | 0.394 | 0.257 | 0.183 | 0.170 | 0.157 |
| | $\sigma$ | 0.228 | 0.132 | 0.051 | 0.079 | 0.075 |
|  | M | 0.312 | 0.235 | 0.172 | 0.148 | 0.138 |
| SCVPM (TMB) | $\mu$ | 29.3 | 4.0 | 3.5 | 7.7 | 8.1 |
| | $\sigma$ | 32.5 | 2.6 | 3.3 | 28.3 | 28.9 |
|  | M | 19.2 | 3.3 | 3.1 | 2.2 | 2.2 |

**Supplementary Table 15.**
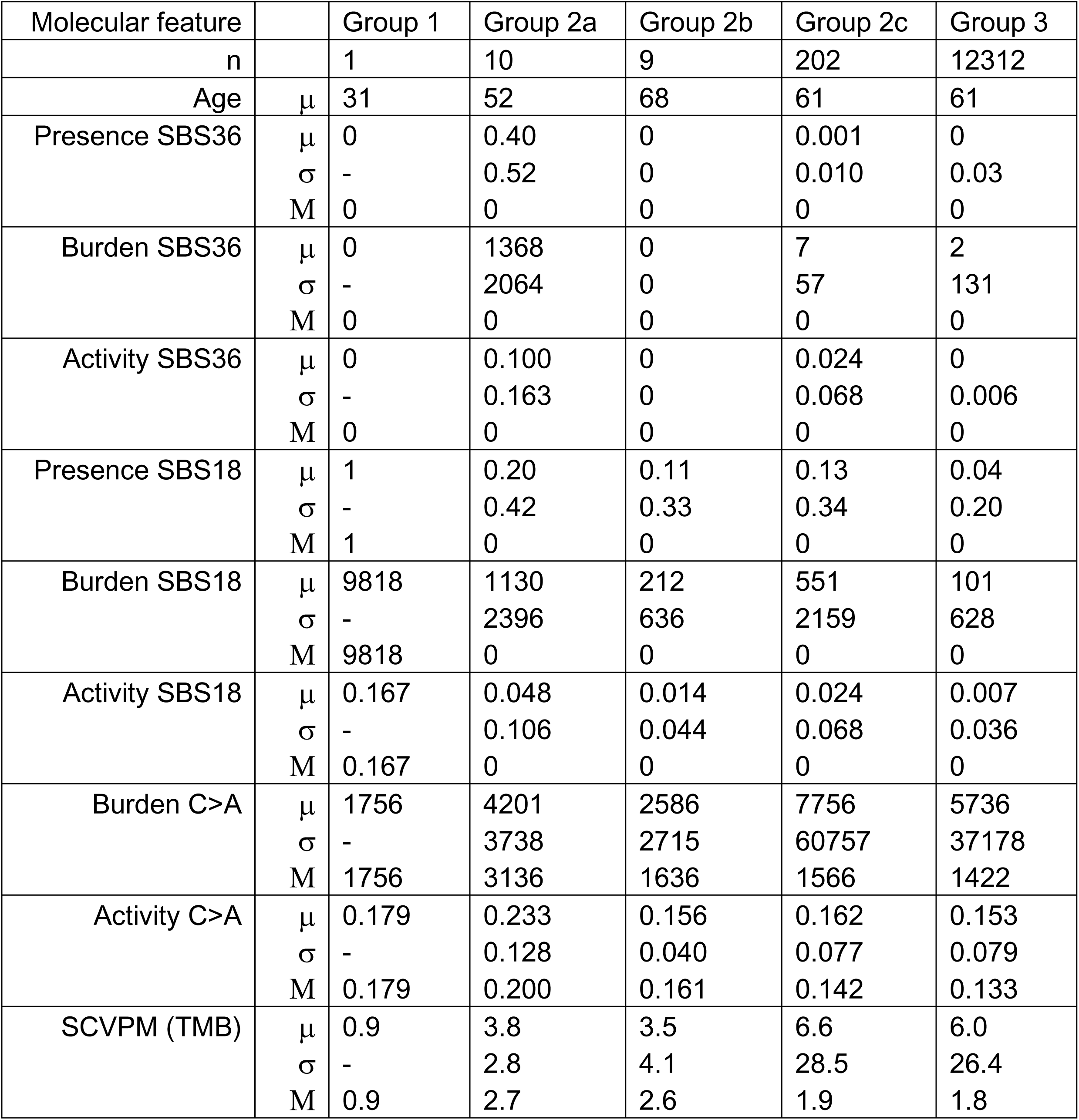
Summary of somatic molecular features of 100kGP CRC-excluded tumours. Legend as in **Supplementary Table 2**.

| Molecular feature |  | Group 1 | Group 2a | Group 2b | Group 2c | Group 3 |
| --- | --- | --- | --- | --- | --- | --- |
| n |  | 1 | 10 | 9 | 202 | 12312 |
| Age | $\mu$ | 31 | 52 | 68 | 61 | 61 |
| Presence SBS36 | $\mu$ | 0 | 0.40 | 0 | 0.001 | 0 |
| | $\sigma$ | - | 0.52 | 0 | 0.010 | 0.03 |
|  | M | 0 | 0 | 0 | 0 | 0 |
| Burden SBS36 | $\mu$ | 0 | 1368 | 0 | 7 | 2 |
| | $\sigma$ | - | 2064 | 0 | 57 | 131 |
|  | M | 0 | 0 | 0 | 0 | 0 |
| Activity SBS36 | $\mu$ | 0 | 0.100 | 0 | 0.024 | 0 |
| | $\sigma$ | - | 0.163 | 0 | 0.068 | 0.006 |
|  | M | 0 | 0 | 0 | 0 | 0 |
| Presence SBS18 | $\mu$ | 1 | 0.20 | 0.11 | 0.13 | 0.04 |
| | $\sigma$ | - | 0.42 | 0.33 | 0.34 | 0.20 |
|  | M | 1 | 0 | 0 | 0 | 0 |
| Burden SBS18 | $\mu$ | 9818 | 1130 | 212 | 551 | 101 |
| | $\sigma$ | - | 2396 | 636 | 2159 | 628 |
|  | M | 9818 | 0 | 0 | 0 | 0 |
| Activity SBS18 | $\mu$ | 0.167 | 0.048 | 0.014 | 0.024 | 0.007 |
| | $\sigma$ | - | 0.106 | 0.044 | 0.068 | 0.036 |
|  | M | 0.167 | 0 | 0 | 0 | 0 |
| Burden C>A | $\mu$ | 1756 | 4201 | 2586 | 7756 | 5736 |
| | $\sigma$ | - | 3738 | 2715 | 60757 | 37178 |
|  | M | 1756 | 3136 | 1636 | 1566 | 1422 |
| Activity C>A | $\mu$ | 0.179 | 0.233 | 0.156 | 0.162 | 0.153 |
| | $\sigma$ | - | 0.128 | 0.040 | 0.077 | 0.079 |
|  | M | 0.179 | 0.200 | 0.161 | 0.142 | 0.133 |
| SCVPM (TMB) | $\mu$ | 0.9 | 3.8 | 3.5 | 6.6 | 6.0 |
| | $\sigma$ | - | 2.8 | 4.1 | 28.5 | 26.4 |
|  | M | 0.9 | 2.7 | 2.6 | 1.9 | 1.8 |

**Supplementary Table 16.** Summary of somatic molecular features of Upper GI cancers. Legend as in **Supplementary Table 2**.

| Molecular feature |  | Group 2c (all MSS) | Group 3 (all) | Group 3 (MSS only) |
| --- | --- | --- | --- | --- |
| n |  | 10 | 371 | 360 |
| Age | $\mu$ | 69 | 67 | 69 |
| Presence SBS36 | $\mu$ | 0 | 0 | 0 |
| | $\sigma$ | 0 | 0 | 0 |
|  | M | 0 | 0 | 0 |
| Burden SBS36 | $\mu$ | 0 | 0 | 0 |
| | $\sigma$ | 0 | 0 | 0 |
|  | M | 0 | 0 | 0 |
| Activity SBS36 | $\mu$ | 0 | 0 | 0 |
| | $\sigma$ | 0 | 0 | 0 |
|  | M | 0 | 0 | 0 |
| Presence SBS18 | $\mu$ | 0.30 | 0.18 | 0.19 |
| | $\sigma$ | 0.48 | 0.39 | 0.40 |
|  | M | 0 | 0 | 0 |
| Burden SBS18 | $\mu$ | 2346 | 726 | 809 |
| | $\sigma$ | 4476 | 1771 | 1943 |
|  | M | 0 | 0 | 0 |
| Activity SBS18 | $\mu$ | 0.078 | 0.031 | 0.032 |
| | $\sigma$ | 0.132 | 0.070 | 0.071 |
|  | M | 0 | 0 | 0 |
| Burden C>A | $\mu$ | 4287 | 2812 | 2527 |
| | $\sigma$ | 3339 | 2712 | 1894 |
|  | M | 2583 | 2136 | 2043 |
| Activity C>A | $\mu$ | 0.191 | 0.144 | 0.146 |
| | $\sigma$ | 0.058 | 0.036 | 0.037 |
|  | M | 0.192 | 0.139 | 0.140 |
| SCVPM (TMB) | $\mu$ | 3.5 | 4.7 | 2.9 |
| | $\sigma$ | 2.5 | 11.0 | 3.2 |
|  | M | 2.8 | 2.4 | 2.3 |

**Supplementary Table 17.** Multiple regression analysis of 100kGP All-cancer and CRC-excluded data sets. Incorporated co-variables were (1) origin from metastasis, (2) older age, (3) male sex, (4) lower tumour purity, (5) higher TMB/SCVPM (unless the outcome), (6) Prior radiotherapy, (7) Prior chemotherapy, (8) FFPE or PCR sequencing library. Note that CRC sequences were all from PCR-free libraries, and thus (8) was only used for All-cancer analysis. The three main tests – germline bi-allelic *MUTYH* mutation carriers versus others (group 1 *vs* groups 2+3), mono-allelic mutation carriers with somatic loss of the wildtype allele (Group 2a *vs* groups 2b/c) & mono-allelic carriers *vs* wildtype (group 2 *vs* group 3) – are shown. ND, not analysed owing to small sizes of some groups, NS, not significant in final regression model at nominal P<0.05. Columns “Other cov” show co-variables remaining in the final model at nominal P<0.05. If the direction of effect of the associated variable is opposite to the listed above, it is shown by a – sign.

| 1 vs 2+3 | All-cancer |  |  |  | CRC-excluded |  |  |  |
| --- | --- | --- | --- | --- | --- | --- | --- | --- |
| Somatic features | $\beta$ | SE | P | Other cov | $\beta$ | SE | P | Other cov |
| Presence SBS36 | 0.599 | 0.245 | 0.015 | -5,-8 |  |  | ND |  |
| Burden SBS36 | 19487 | 7972 | 0.015 |  |  |  | ND |  |
| Activity SBS36 |  |  | NS |  |  |  | ND |  |
| Presence SBS18 | 0.533 | 0.024 | 0.025 | 2,3,-5,-6,7,-8 |  |  | ND |  |
| Burden SBS18 |  |  | NS |  |  |  | ND |  |
| Activity SBS18 | 0.158 | 0.077 | 0.040 | 2,3,-5,-6,7,-5,-8 |  |  | ND |  |
| Burden C>A |  |  | NS |  |  |  | ND |  |
| Activity C>A | 0.236 | 0.103 | 0.022 | 2,3,-5,-6,7,-5,-8 |  |  | ND |  |
| TMB |  |  | NS |  |  |  | ND |  |

| 2a vs 2b/c | All-cancer |  |  |  | CRC-excluded |  |  |  |
| --- | --- | --- | --- | --- | --- | --- | --- | --- |
| Somatic features | $\beta$ | SE | P | Other cov | $\beta$ | SE | P | Other cov |
| Presence SBS36 | 0.405 | 0.007 | $6.90 \times 10^{-3}$ | | 0.386 | 0.164 | 0.019 | |
| Burden SBS36 | 2134 | 1052 | 0.043 |  | 1361 | 653 | 0.038 |  |
| Activity SBS36 | 0.121 | 0.053 | 0.024 |  |  |  | NS |  |
| Presence SBS18 |  |  | NS |  |  |  | NS |  |
| Burden SBS18 |  |  | NS |  |  |  | NS |  |
| Activity SBS18 |  |  | NS |  |  |  | NS |  |
| Burden C>A |  |  | NS |  |  |  | NS |  |
| Activity C>A | 0.096 | 0.014 | $3.63 \times 10^{-3}$ | -1,2 | 0.084 | 0.034 | 0.013 | -1,2,5 |
| TMB |  |  | NS |  |  |  | NS |  |

| 2 vs 3 | All-cancer |  |  |  | CRC-excluded |  |  |  |
| --- | --- | --- | --- | --- | --- | --- | --- | --- |
| Somatic features | $\beta$ | SE | P | Other cov | $\beta$ | SE | P | Other cov |
| Presence SBS36 | 0.028 | 0.010 | 0.006 | -5,-8 | 0.031 | 0.012 | $8.59 \times 10^{-3}$ | -5,-8 |
| Burden SBS36 |  |  | NS |  |  |  | NS |  |
| Activity SBS36 | $6.06 \times 10^{-3}$ | $2.77 \times 10^{-3}$ | 0.028 | -5,-8 | $5.47 \times 10^{-3}$ | $2.68 \times 10^{-3}$ | 0.041 | -8 |
| Presence SBS18 | 0.116 | 0.024 | $1.56 \times 10^{-6}$ | 2,-5,-8 | 0.088 | 0.023 | $1.19 \times 10^{-4}$ | -1,2,-3,-5,-8 |
| Burden SBS18 | 732 | 175 | $2.99 \times 10^{-5}$ | 2,-5,-8 | 462 | 177143 | $1.22 \times 10^{-3}$ | -1,2,3,-5,7,-8 |
| Activity SBS18 | 0.029 | 0.006 | $9.68 \times 10^{-7}$ | 2,-5,-8 | 0.018 | 0.005 | $1.08 \times 10^{-5}$ | -1,2,-5,-8 |
| Burden C>A |  |  | NS |  |  |  | NS |  |
| Activity C>A | 0.017 | 0.005 | $6.84 \times 10^{-4}$ | -1,2,4,5,-6,7,-8 | 0.010 | 0.005 | 0.042 | -1,2,4,5,-6,7,-8 |
| TMB |  |  | NS |  |  |  | NS |  |

**Supplementary Table 18.** Pairwise associations in univariable analyses between germline MUTYH genotypes and molecular features in upper gastrointestinal, breast, prostate and endometrial cancers from 100kGP. Upper GI cancers included, cholangiocarcinomas (n=33), gastric carcinomas (n=81), pancreatic carcinomas (n=125), hepatocellular carcinomas (n=24), oesophageal adenocarcinomas (n=96), small intestinal adenocarcarcinomas (n=8) and others (n=16). For binary variables, P are values from χ^2^ or Fisher’s exact tests, whereas for associations involving quantitative variables (burden and activity), P values are from Wilcoxon tests. Effect size metrics (i.e. βs (log(ORs)), standard errors (SE) and corresponding P_regress_ values) are estimated from univariable regression with robust variances. ND shows tests not performed owing to small sample numbers in one or more groups. No bi-allelic germline mutation carriers were present in any of the three cancer types. No prostate cancer showed LOH at the *MUTYH* locus. No upper GI cancer was from group 2a or 2b. SBS36 was very rare (<0.2% presence) or absent in all four cancer types. SBS18 was present in 19% upper GI, 4% breast, 1% prostate and 17% endometrial cancers. It is likely that the low SBS18 prevalence in breast and prostate tumours, relatively small sample size in upper GI, prostate and endometrial cancers, and heterogeneity of upper GI cancer types all hampered the power of the statistical tests, especially the regression analyses, given that the frequency of mono-allelic mutation carriers is only ∼2.0%. The non-parametric and categorical tests are, however, consistent with positive effects of loss of the germline wildtype allele (group 2a) and of the mono-allelic germline genotype (group 2) on SBS36 and SBS18 respectively.

| MUTYH Groups | Somatic features | Upper GI cancer (n=383) |  |  |  | Breast cancer (n=2,981) |  |  |  | Prostate cancer (n=574) |  |  |  | Endometrial cancer (n=835) |  |  |  |
| --- | --- | --- | --- | --- | --- | --- | --- | --- | --- | --- | --- | --- | --- | --- | --- | --- | --- |
| | | P | $\beta$ | SE | $P_{\text{regress}}$ | P | $\beta$ | SE | $P_{\text{regress}}$ | P | $\beta$ | SE | $P_{\text{regress}}$ | P | $\beta$ | SE | $P_{\text{regress}}$ |
| 2a vs 2b/c | Presence SBS36 | ND | ND | ND | ND | 0.083 | ND | ND | ND | ND | ND | ND | ND | 0.11 | ND | ND | ND |
|  | Burden SBS36 | ND | ND | ND | ND | 2.71x10<sup>-6</sup> | ND | ND | ND | ND | ND | ND | ND | 6.15x10<sup>-5</sup> | ND | ND | ND |
|  | Activity SBS36 | ND | ND | ND | ND | 2.71x10<sup>-6</sup> | ND | ND | ND | ND | ND | ND | ND | 6.15x10<sup>-5</sup> | ND | ND | ND |
|  | Presence SBS18 | ND | ND | ND | ND | 1 | ND | ND | ND | ND | ND | ND | ND | 1 | ND | ND | ND |
|  | Burden SBS18 | ND | ND | ND | ND | 0.62 | ND | ND | ND | ND | ND | ND | ND | 0.70 | ND | ND | ND |
|  | Activity SBS18 | ND | ND | ND | ND | 0.62 | ND | ND | ND | ND | ND | ND | ND | 0.70 | ND | ND | ND |
|  | Burden C>A | ND | ND | ND | ND | 0.13 | ND | ND | ND | ND | ND | ND | ND | 0.74 | ND | ND | ND |
|  | Activity C>A | ND | ND | ND | ND | 0.032 | ND | ND | ND | ND | ND | ND | ND | 0.21 | ND | ND | ND |
|  | TMB | ND | ND | ND | ND | 0.52 | ND | ND | ND | ND | ND | ND | ND | 0.78 | ND | ND | ND |
| 2 vs 3 | Presence SBS36 | ND | ND | ND | ND | 0.19 | 0.019 | 0.021 | 0.360 | 0.035 | ND | ND | ND | 0.046 | ND | ND | ND |
|  | Burden SBS36 | ND | ND | ND | ND | 0.003 | 0.113 | 0.115 | 0.326 | 6.55x10<sup>-14</sup> | ND | ND | ND | 5.93x10<sup>-11</sup> | ND | ND | ND |
|  | Activity SBS36 | ND | ND | ND | ND | 0.003 | 0.010 | 0.010 | 0.326 | 6.55x10<sup>-14</sup> | ND | ND | ND | 5.93x10<sup>-11</sup> | ND | ND | ND |
|  | Presence SBS18 | 0.535 | 0.183 | 0.020 | 0.449 | 0.013 | 0.091 | 0.048 | 0.061 | 0.003 | 0.196 | 0.133 | 0.141 | 0.55 | 0.071 | 0.097 | 0.465 |
|  | Burden SBS18 | 0.260 | 1620 | 1418 | 0.254 | 7.06x10<sup>-4</sup> | 169 | 95 | 0.076 | 1.47x10<sup>-13</sup> | 134 | 93 | 0.151 | 0.38 | 101 | 153 | 0.511 |
|  | Activity SBS18 | 0.212 | 0.046 | 0.042 | 0.266 | 6.56x10<sup>-4</sup> | 0.019 | 0.010 | 0.060 | 1.47x10<sup>-13</sup> | 0.023 | 0.015 | 0.143 | 0.33 | 0.016 | 0.018 | 0.376 |
|  | Burden C>A | 0.195 | 1474 | 1065 | 0.167 | 0.18 | 46 | 143 | 0.745 | 0.55 | -589 | 249 | 0.018 | 0.60 | 18851 | 45200 | 0.677 |
|  | Activity C>A | 0.003 | 0.048 | 0.018 | 0.010 | 0.001 | 0.021 | 0.008 | 0.009 | 0.18 | 0.008 | 0.007 | 0.201 | 0.067 | 0.024 | 0.022 | 0.260 |
|  | TMB | 0.545 | -1.25 | 0.978 | 0.202 | 0.86 | -0.49 | 0.27 | 0.072 | 0.37 | -0.60 | 0.246 | 0.016 | 0.89 | 8.04 | 20.1 | 0.690 |

**Supplementary Table 19.** Multiple regression analysis of Upper GI cancers. Incorporated co-variables were (1) origin from metastasis, (2) older age, (3) male sex, (4) lower tumour purity, (5) higher TMB/SCVPM (unless the outcome), (6) Prior radiotherapy, (7) Prior chemotherapy, (8) FFPE or PCR sequencing library, (9) MSI. Legend is otherwise as per **Supplementary Table 16**. If the direction of effect of the associated variable is opposite to the listed above, it is shown by a – sign.

| 2 vs 3 | Upper GI cancers |  |  |  |
| --- | --- | --- | --- | --- |
| Somatic features | $\beta$ | SE | P | Other cov |
| Presence SBS18 | 0.116 | 0.024 | NS |  |
| Burden SBS18 | 732 | 175 | NS |  |
| Activity SBS18 | 0.029 | 0.006 | NS |  |
| Burden C>A | 1885 | 827 | 0.023 | 3, 5, -4 |
| Activity C>A | 0.051 | 0.018 | 0.004 | 1, 9, -4 |
| TMB |  |  | NS |  |

**Supplementary Table 20.** Associations between germline MUTYH genotypes and All-cancer risk in 100kGP. Two-tailed Fisher’s exact tests. To assess the overall cancer risk associated with germline *MUTYH* mutations, we studied 11,318 unrelated participants of European genetic ancestry with a malignant neoplasm in the 100kGP cancer domain. There was no specific enrichment of cases with colorectal polyposis. Germline bi-allelic mutation of *MUTYH* was very rare (frequency <0.05%), principally occurring in participants with colorectal cancer (compared with the control data set, OR=6.0, 95%CI 0.43-82.57, *P=*0.10), but occasionally present in other cancer cases. Considering all tumour types together, there was no significant enrichment of bi-allelic germline *MUTYH* mutations in cancer cases compared to the set of cancer-free controls (OR=1.90, 95%CI 0.22-22.77; *P=*0.66, *P_adj_=*1.00).

| Germline *MUTYH* | OR (95% CI) | P | P<sub>adj</sub> |
| --- | --- | --- | --- |
| Bi-allelic mutation | 1.90 (0.22-22.77) | 0.66 | 1.00 |
| Mono-allelic mutation | 1.01 (0.83-1.21) | 0.96 | 1.00 |
| Mono-allelic p.Gly396Asp | 0.92 (0.74-1.15) | 0.47 | 1.00 |
| Mono-allelic p.Tyr179Cys | 1.41 (0.92-2.14) | 0.10 | 0.50 |
| Mono-allelic PTV | 1.09 (0.30-3.78) | 1.00 | 1.00 |

**Supplementary Table 21.** Associations between various germline MUTYH genotypes and All-cancer risk in UKB. Two-tailed Fisher’s exact tests. We used WES data from 12,694 unrelated cases of European genetic ancestry with a malignant neoplasm and 28,939 European unrelated, cancer-free controls from UKBiobank. Considering all tumour types together, there was a significant enrichment of bi-allelic germline *MUTYH* mutations in cancer cases compared to the set of controls (OR=15.97, 95%CI 2.05-717; *P=*1.44x10^-3^). However, this was largely driven by six cases (0.05%) with CRC, and one control (0.004%) with presumed bi-allelic germline *MUTYH* mutations, driving a significant overall association with cancer risk (see also **Supplementary Table 13**).

| Germline *MUTYH* | OR (95% CI) | P | P<sub>adj</sub> |
| --- | --- | --- | --- |
| Bi-allelic mutation | 15.97 (2.05-717.19) | $1.44 \times 10^{-3}$ | 0.01 |
| Mono-allelic mutation | 1.01 (0.86-1.18) | 0.90 | 0.93 |
| Mono-allelic p.Gly396Asp | 1.02 (0.84-1.24) | 0.85 | 0.93 |
| Mono-allelic p.Tyr179Cys | 0.97 (0.69-1.36) | 0.93 | 0.93 |
| Mono-allelic PTV | 1.74 (0.78-3.82) | 0.16 | 0.40 |

**Supplementary Table 22.** Associations between bi-allelic germline MUTYH mutations and risk of individual cancer types in 100kGP. Two-tailed Fisher’s exact tests. We did not find significantly elevated levels of germline bi-allelic mutations in any of the individual cancer types before or after correcting for multiple testing. Note that the numbers of cases in 100kGP data may not be shown exactly if they lie between 1 and 5, for reasons of patient confidentiality. Germline bi-allelic loss of *MUTYH* has specifically been suggested to predispose to gynaecological cancers (including ovarian and endometrial) and breast cancers (Paller et al., 2024; Win et al., 2016), but we identified no participants with these cancer types who had germline bi-allelic *MUTYH* inactivation.

| Cancer type | Bi-allelic mutation | Total | OR (95% CI) | P | Padj |
| --- | --- | --- | --- | --- | --- |
| Bones & joints | 0 | 227 | 0 (0-334.51) | 1.00 | 1.00 |
| Brain & other nervous system | 0 | 514 | 0 (0-148.31) | 1.00 | 1.00 |
| Breast | 0 | 2463 | 0 (0-31.03) | 1.00 | 1.00 |
| Colon & rectum | <5 | 2395 | 6.0 (0.43-82.57) | 0.10 | 1.00 |
| Endocrine system | 0 | 72 | 0 (0-1031.6) | 1.00 | 1.00 |
| Eye & orbit | 0 | 18 | 0 (0-4588.58) | 1.00 | 1.00 |
| Female genital system | 0 | 1258 | 0 (0-61.23) | 1.00 | 1.00 |
| Leukaemia | 0 | 307 | 0 (0-245.47) | 1.00 | 1.00 |
| Liver & intrahepatic bile duct | 0 | 289 | 0 (0-264.51) | 1.00 | 1.00 |
| Lymphoma | 0 | 114 | 0 (0-663.45) | 1.00 | 1.00 |
| Male genital system | <5 | 794 | 9.08 (0.15-174) | 0.15 | 1.00 |
| Mesothelioma | 0 | <5 | 0 (0-Inf) | 1.00 | 1.00 |
| Miscellaneous | 0 | 91 | 0 (0-842.02) | 1.00 | 1.00 |
| Myeloma | 0 | 99 | 0 (0-799.37) | 1.00 | 1.00 |
| Oral cavity & pharynx | 0 | 232 | 0 (0-334.51) | 1.00 | 1.00 |
| Respiratory system | 0 | 1362 | 0 (0-56.12) | 1.00 | 1.00 |
| Skin (excluding BCC & SCC) | 0 | 1205 | 0 (0-63.35) | 1.00 | 1.00 |
| Soft tissue including heart | 0 | 513 | 0 (0-147.72) | 1.00 | 1.00 |
| Upper gastrointestinal | 0 | 314 | 0 (0-246.27) | 1.00 | 1.00 |
| Urinary system | 0 | 1603 | 0 (0-47.64) | 1.00 | 1.00 |

**Supplementary Table 23.** Associations between bi-allelic germline MUTYH mutations and risk of individual cancer types in UKB. Two-tailed Fisher’s exact tests. With the exception of CRC, there was no evidence that bi-allelic germline MUYTH carriers were at increased risk of any specific cancer type.

| Cancer type | Bi-allelic mutation | Total | OR | P | Padj |
| --- | --- | --- | --- | --- | --- |
| Bones & joints | 0 | 21 | 0 [0-Inf] | 1.00 | 1.00 |
| Brain & other nervous system | 0 | 109 | 0 [0-8772.74] | 1.00 | 1.00 |
| Breast | 1 | 2763 | 10.49 [0.13-817.67] | 0.17 | 1.00 |
| Colon & rectum | 6 | 1067 | 164.37 [19.9-7159] | 0.00 | 0.00 |
| Endocrine system | 0 | 124 | 0 [0-8129.18] | 1.00 | 1.00 |
| Eye & orbit | 0 | 39 | 0 [0-16384] | 1.00 | 1.00 |
| Female genital system | 0 | 819 | 0 [0-1349.21] | 1.00 | 1.00 |
| Leukaemia | 0 | 220 | 0 [0-4729.22] | 1.00 | 1.00 |
| Liver & intrahepatic bile duct | 0 | 225 | 0 [0-4649.84] | 1.00 | 1.00 |
| Lymphoma | 1 | 519 | 56.27 [0.72-4229] | 0.03 | 0.35 |
| Male genital system | 0 | 1794 | 0 [0-620] | 1.00 | 1.00 |
| Mesothelioma | 0 | 40 | 0 [0-16384] | 1.00 | 1.00 |
| Miscellaneous | 0 | 14 | 0 [0-Inf] | 1.00 | 1.00 |
| Myeloma | 0 | 121 | 0 [0-8011.63] | 1.00 | 1.00 |
| Oral cavity & pharynx | 0 | 183 | 0 [0-5559.68] | 1.00 | 1.00 |
| Respiratory system | 0 | 452 | 0 [0-2396.91] | 1.00 | 1.00 |
| Skin (excluding BCC & SCC) | 1 | 4506 | 6.41 [0.08-501.42] | 0.25 | 1.00 |
| Soft tissue including heart | 0 | 109 | 0 [0-8986.17] | 1.00 | 1.00 |
| Upper gastrointestinal | 0 | 264 | 0 [0-4039.61] | 1.00 | 1.00 |
| Urinary system | 0 | 460 | 0 [0-2376.35] | 1.00 | 1.00 |

**Supplementary Table 24.** Associations between mono-allelic germline MUTYH mutations and risk of various non-CRC cancers in 100kGP. Results are from two-tailed Fisher’s exact tests. Five cancer (sub-) types studied by Barreiro et al are shown separately, indicated by *. Mono-allelic pathogenic germline MUTYH mutations (group 2) were present in 211 (1.80%) cases, a very similar frequency to that in controls (OR=1.01, 95% CI 0.83-1.21; P=0.96, Padj=1.00). Mono-allelic mutations were generally no more common in any of the other 19 ICD cancer types compared to controls, albeit with limited evidence of an increased risk of gynaecological cancer (OR=1.47, 95%CI 0.99-2.12; P=0.04, P_adj_=0.58). There was no association between mono-allelic germline MUTYH mutation and CRC-excluded cancer risk (OR=0.907 95%CI 0.755-1.090). + exact counts available through 100kGP.

| Cancer type | Mono-allelic mutation * | Total | OR [95% CI] | P | Padj |
| --- | --- | --- | --- | --- | --- |
| Bones & joints | <5 | 227 | 0.47 [0.06-1.74] | 0.45 | 1.00 |
| Brain & other nervous system | 8 | 514 | 0.84 [0.36-1.69] | 0.74 | 1.00 |
| Breast | 45 | 2463 | 0.98 [0.70-1.36] | 1.00 | 1.00 |
| Endocrine system | <5 | 72 | 0.75 [0.02-4.33] | 1.00 | 1.00 |
| Eye & orbit | 0 | 18 | 0 [0-12.11] | 1.00 | 1.00 |
| Female genital system | 34 | 1258 | 1.47 [0.99-2.12] | 0.04 | 0.58 |
| Leukaemia | <5 | 307 | 0.17 [0-0.98] | 0.05 | 0.58 |
| Liver & intrahepatic bile duct | 5 | 289 | 0.93 [0.30-2.23] | 1.00 | 1.00 |
| Lymphoma | <5 | 114 | 0.47 [0.01-2.69] | 0.73 | 1.00 |
| Male genital system | 18 | 794 | 1.23 [0.71-1.99] | 0.42 | 1.00 |
| Mesothelioma | 0 | <5 | 0 [0-1999.47] | 1.00 | 1.00 |
| Miscellaneous | <5 | 91 | 0.59 [0.01-3.39] | 1.00 | 1.00 |
| Myeloma | <5 | 99 | 2.23 [0.59-5.96] | 0.11 | 0.73 |
| Oral cavity & pharynx | 7 | 232 | 1.65 [0.65-3.50] | 0.21 | 0.73 |
| Respiratory system | 25 | 1362 | 0.99 [0.63-1.50] | 1.00 | 1.00 |
| Skin (excluding BCC & SCC) | 22 | 1205 | 0.98 [0.60-1.53] | 1.00 | 1.00 |
| Soft tissue including heart | 5 | 513 | 0.52 [0.17-1.24] | 0.18 | 0.73 |
| Upper gastrointestinal | 9 | 314 | 1.56 [0.70-3.05] | 0.20 | 0.73 |
| Urinary system | 30 | 1603 | 1.01 [0.67-1.48] | 0.92 | 1.00 |
| Oesophageal adenoca. * | 0 | 18 | 0 [0-12.11] | 1.00 | 1.00 |
| Prostate adenocarcinoma * | 10 | 410 | 1.32 [0.62-2.50] | 0.35 | 1.00 |
| Renal clear cell * | 12 | 607 | 1.07 [0.54-1.91] | 0.76 | 1.00 |
| Sarcoma * | 8 | 742 | 0.58 [0.25-1.16] | 0.16 | 0.73 |

**Supplementary Table 25.** Associations between mono-allelic germline MUTYH mutations and risk of various non-CRC cancers in UKB. Two-tailed Fisher’s exact tests. Five cancer (sub-)types studied by Barreiro et al are shown separately, indicated by *. Mono-allelic pathogenic germline MUTYH mutations (group 2) were present in 239 (1.88%) UKB cases, a very similar frequency to that in controls (OR=0.99, 95% CI 0.84-1.15; P=0.88, Padj=0.93). Overall, the frequency of mono-allelic mutations was not significantly elevated in any of the 20 cancer types compared to the controls in UKB, including gynaecological cancers. There was no association between mono-allelic germline MUTYH mutation and CRC-excluded cancer risk (OR=0.972 95%CI 0.824-1.146; in meta-analysis with 100kGP data, OR=0.941, 95%CI 0.830-1.072).

| Cancer type | Mono-allelic mutation | Total | OR | P | Padj |
| --- | --- | --- | --- | --- | --- |
| Bones & joints | 1 | 21 | 2.57 [0.06-16.09] | 0.33 | 0.66 |
| Brain & other nervous system | 0 | 109 | 0 [0-1.77] | 0.28 | 0.66 |
| Breast | 55 | 2763 | 1.04 [0.77-1.38] | 0.77 | 1.00 |
| Endocrine system | 6 | 124 | 2.61 [0.93-5.89] | 0.03 | 0.55 |
| Eye & orbit | 0 | 39 | 0 [0-5.11] | 1.00 | 1.00 |
| Female genital system | 17 | 819 | 1.09 [0.63-1.77] | 0.70 | 0.97 |
| Leukaemia | 3 | 220 | 0.71 [0.14-2.11] | 0.80 | 1.00 |
| Liver & intrahepatic bile duct | 4 | 225 | 0.93 [0.25-2.43] | 1.00 | 1.00 |
| Lymphoma | 13 | 519 | 1.32 [0.69-2.3] | 0.33 | 0.66 |
| Male genital system | 24 | 1794 | 0.70 [0.44-1.05] | 0.09 | 0.55 |
| Mesothelioma | 1 | 40 | 1.32 [0.03-7.80] | 0.54 | 0.79 |
| Miscellaneous | 0 | 14 | 0 [0-15.51] | 1.00 | 1.00 |
| Myeloma | 0 | 121 | 0 [0-1.59] | 0.18 | 0.60 |
| Oral cavity & pharynx | 1 | 183 | 0.28 [0.01-1.60] | 0.27 | 0.66 |
| Respiratory system | 8 | 452 | 0.92 [0.39-1.85] | 1.00 | 1.00 |
| Skin excluding BCC & SCC | 77 | 4506 | 0.89 [0.69-1.14] | 0.38 | 0.66 |
| Soft tissue including heart | 3 | 109 | 1.45 [0.29-4.38] | 0.47 | 0.73 |
| Upper gastrointestinal | 7 | 264 | 1.40 [0.55-2.94] | 0.36 | 0.66 |
| Urinary system | 11 | 460 | 1.26 [0.62-2.29] | 0.39 | 0.66 |
| Adrenocortical carcinoma * | 0 | 2 | 0 [0-273.63] | 1.00 | 1.00 |
| Oesophageal * | 5 | 135 | 1.97 [0.63-4.75] | 0.19 | 0.60 |
| Prostate * | 21 | 1634 | 0.67 [0.41-1.04] | 0.07 | 0.55 |
| Renal * | 8 | 215 | 1.98 [0.84-4.01] | 0.07 | 0.55 |
| Sarcoma * | 3 | 66 | 2.44 [0.49-7.51] | 0.13 | 0.58 |

**Supplementary Table 26.** Germline de novo mutations in offspring of carriers of mono-allelic MUTYH mutations. The table entries are the numbers of *de novo* mutations (DNMs) identified by 100kGP in offspring for which a single parent is a monby 100kGP.o-allelic *MUTYH* mutation carrier compared offspring who have a single parent who is a carrier of bi-allelic germline *MUTYH* mutations, and offspring whose parents have no germline *MUTYH* mutation. Data from stringent and non-stringent DNM calling pipelines are provided, as defined by 100kGP (see link in **Methods**). *P_stringent_* mono-allelic *v* wildtype = 0.55, t test; *P_non-stringent_* mono-allelic *v* wildtype = 0.016, t test). Stringent calls are recommended for use

|  | Non-stringent |  |  | Stringent |  |  |
| --- | --- | --- | --- | --- | --- | --- |
| MUTYH genotype | Mean | SD | Median | Mean | SD | Median |
| Bi-allelic (n=2) | 910 | 134 | 910 | 63 | 17 | 63 |
| Mono-allelic (n=206) | 1033 | 548 | 948 | 69 | 15 | 69 |
| Wildtype (n=11,308) | 980 | 306 | 915 | 70 | 24 | 69 |

**Supplementary Table 27.** Germline mono-allelic MBD4 mutations, mutational processes in 100kGP cancers, and CRC risk. (a) Summary of P values for tests of association between mono-allelic MBD4 mutation carriers and each of the four measures of C>T mutation in 100kGP cancers. Owing to limited data, and hence some uncertainty, regarding the effects of missense MBD4 mutations, only protein-truncating mutations (stop-gained, frameshift or pathogenic splice, supported by Splice AI and Clinvar) were regarded as pathogenic. Owing to the presence of a short repeat in MBD4 that is prone to background slippage in MSI+ cancers, the analyses were undertaken in MSS cancers only. In all instances with a significant association, the MBD4 carriers showed higher measures than the wildtype comparison group. Single variable analyses used Wilcoxon test. Multivariable analysis used linear regression with robust variances, incorporating age, sex, tumour purity, SCVPM and prior therapy as co-variables. The results suggest, plausibly, that CRC-only analyses are underpowered relative to the other, larger data sets.

| Cancer group | Statistical analysis | SBS1 burden | SBS1 activity | C>T burden | C>T activity |
| --- | --- | --- | --- | --- | --- |
| CRC | Univariable | >0.05 | >0.05 | >0.05 | 0.0810 |
| N=2,039 (8 carriers) | Multivariable | >0.05 | >0.05 | >0.05 | >0.05 |
| CRC-excluded | Univariable | 0.0132 | 0.00584 | >0.05 | >0.05 |
| N=12,172 (6 carriers) | Multivariable | >0.05 | >0.05 | >0.05 | >0.05 |
| All cancer | Univariable | 2.73x10<sup>-5</sup> | 3.95x10<sup>-5</sup> | >0.05 | 0.0175 |
| N=14,211 (14 carriers) | Multivariable | 1.43x10<sup>-2</sup> | 9.55x10<sup>-3</sup> | 2.01x10<sup>-2</sup> | >0.05 |

(b) Association between mono-allelic germline *MBD4* mutation and CRC risk in 100kGP and UKB.
| Study | N mono-allelic cases/total | N mono-allelic controls/total | OR | 95%CI | P |
| --- | --- | --- | --- | --- | --- |
| 100kGP + CORGI | 9/2,985 | 15/14,441 | 2.908 | 1.272-6.652 | 0.028 |
| UK Biobank WES | 1/1,224 | 7/9,599 | 1.121 | 0.138-9.121 | 1.000 |
| UK Biobank WGS | 10/4,834 | 281/142,813 | 1.051 | 0.559-1.977 | 0.966 |
| Meta-analysis |  |  | 1.75 | 1.09-2.81 | 0.020 |

The forest plot shows meta-analysis results with inverse variance weighting. $P_{\text{het}}=0.032$ , $I^2=71.0\%$

## References

1. Garutti M, Foffano L, Mazzeo R, Michelotti A, Da Ros L, Viel A, et al. Hereditary Cancer Syndromes: A Comprehensive Review with a Visual Tool. Genes (Basel). 2023;14(5). Epub 20230430. doi: 10.3390/genes14051025. PubMed PMID: 37239385; PubMed Central PMCID: PMCPMC10218093.

2. Howlett NG, Taniguchi T, Olson S, Cox B, Waisfisz Q, De Die-Smulders C, et al. Biallelic inactivation of BRCA2 in Fanconi anemia. Science. 2002;297(5581):606–9. Epub 20020613. doi: 10.1126/science.1073834. PubMed PMID: 12065746.

3. Wang Q, Lasset C, Desseigne F, Frappaz D, Bergeron C, Navarro C, et al. Neurofibromatosis and early onset of cancers in hMLH1-deficient children. Cancer Res. 1999;59(2):294–7. PubMed PMID: 9927034.

4. Renwick A, Thompson D, Seal S, Kelly P, Chagtai T, Ahmed M, et al. ATM mutations that cause ataxia-telangiectasia are breast cancer susceptibility alleles. Nat Genet. 2006;38(8):873–5. Epub 20060709. doi: 10.1038/ng1837. PubMed PMID: 16832357.

5. Rahman N, Scott RH. Cancer genes associated with phenotypes in monoallelic and biallelic mutation carriers: new lessons from old players. Hum Mol Genet. 2007;16 Spec No 1:R60-6. doi: 10.1093/hmg/ddm026. PubMed PMID: 17613548.

6. Knudson AG. Hereditary cancer: two hits revisited. J Cancer Res Clin Oncol. 1996;122(3):135–40. doi: 10.1007/bf01366952. PubMed PMID: 8601560.

7. Nielsen M, Infante E, Brand R. MUTYH Polyposis. In: Adam MP, Feldman J, Mirzaa GM, Pagon RA, Wallace SE, Amemiya A, editors. GeneReviews. Seattle (WA): University of Washington, Seattle; 1993.

8. Al-Tassan N, Chmiel NH, Maynard J, Fleming N, Livingston AL, Williams GT, et al. Inherited variants of MYH associated with somatic G:C-->T:A mutations in colorectal tumors. Nat Genet. 2002;30(2):227–32. Epub 20020130. doi: 10.1038/ng828. PubMed PMID: 11818965.

9. Cheadle JP, Sampson JR. MUTYH-associated polyposis--from defect in base excision repair to clinical genetic testing. DNA Repair (Amst). 2007;6(3):274–9. Epub 20061211. doi: 10.1016/j.dnarep.2006.11.001. PubMed PMID: 17161978.

10. Pilati C, Shinde J, Alexandrov LB, Assié G, André T, Hélias-Rodzewicz Z, et al. Mutational signature analysis identifies MUTYH deficiency in colorectal cancers and adrenocortical carcinomas. J Pathol. 2017;242(1):10–5. Epub 20170329. doi: 10.1002/path.4880. PubMed PMID: 28127763.

11. Viel A, Bruselles A, Meccia E, Fornasarig M, Quaia M, Canzonieri V, et al. A Specific Mutational Signature Associated with DNA 8-Oxoguanine Persistence in MUTYH-defective Colorectal Cancer. EBioMedicine. 2017;20:39–49. Epub 20170413. doi: 10.1016/j.ebiom.2017.04.022. PubMed PMID: 28551381; PubMed Central PMCID: PMCPMC5478212.

12. Georgeson P, Pope BJ, Rosty C, Clendenning M, Mahmood K, Joo JE, et al. Evaluating the utility of tumour mutational signatures for identifying hereditary colorectal cancer and polyposis syndrome carriers. Gut. 2021;70(11):2138–49. Epub 20210107. doi: 10.1136/gutjnl-2019-320462. PubMed PMID: 33414168; PubMed Central PMCID: PMCPMC8260632.

13. Halford SE, Rowan AJ, Lipton L, Sieber OM, Pack K, Thomas HJ, et al. Germline mutations but not somatic changes at the MYH locus contribute to the pathogenesis of unselected colorectal cancers. Am J Pathol. 2003;162(5):1545–8. doi: 10.1016/s0002-9440(10)64288-5. PubMed PMID: 12707038; PubMed Central PMCID: PMCPMC1851182.

14. Theodoratou E, Campbell H, Tenesa A, Houlston R, Webb E, Lubbe S, et al. A large-scale meta-analysis to refine colorectal cancer risk estimates associated with MUTYH variants. British journal of cancer. 2010;103(12):1875–84. Epub 20101109. doi: 10.1038/sj.bjc.6605966. PubMed PMID: 21063410; PubMed Central PMCID: PMCPMC3008602.

15. Paller CJ, Tukachinsky H, Maertens A, Decker B, Sampson JR, Cheadle JP, et al. Pan-Cancer Interrogation of MUTYH Variants Reveals Biallelic Inactivation and Defective Base Excision Repair Across a Spectrum of Solid Tumors. JCO Precis Oncol. 2024;8:e2300251. doi: 10.1200/po.23.00251. PubMed PMID: 38394468; PubMed Central PMCID: PMCPMC10901435.

16. Fernandez-Rozadilla C, Timofeeva M, Chen Z, Law P, Thomas M, Schmit S, et al. Deciphering colorectal cancer genetics through multi-omic analysis of 100,204 cases and 154,587 controls of European and east Asian ancestries. Nat Genet. 2023;55(1):89–99. Epub 20221220. doi: 10.1038/s41588-022-01222-9. PubMed PMID: 36539618; PubMed Central PMCID: PMCPMC10094749.

17. Win AK, Reece JC, Dowty JG, Buchanan DD, Clendenning M, Rosty C, et al. Risk of extracolonic cancers for people with biallelic and monoallelic mutations in MUTYH. International journal of cancer. 2016;139(7):1557–63. Epub 2016/05/20. doi: 10.1002/ijc.30197. PubMed PMID: 27194394; PubMed Central PMCID: PMCPMC5094810.

18. Barreiro RAS, Sabbaga J, Rossi BM, Achatz MIW, Bettoni F, Camargo AA, et al. Monoallelic deleterious MUTYH germline variants as a driver for tumorigenesis. J Pathol. 2022;256(2):214–22. Epub 20211123. doi: 10.1002/path.5829. PubMed PMID: 34816434.

19. Weinstein JN, Collisson EA, Mills GB, Shaw KR, Ozenberger BA, Ellrott K, et al. The Cancer Genome Atlas Pan-Cancer analysis project. Nat Genet. 2013;45(10):1113–20. doi: 10.1038/ng.2764. PubMed PMID: 24071849; PubMed Central PMCID: PMCPMC3919969.

20. Karczewski KJ, Francioli LC, Tiao G, Cummings BB, Alföldi J, Wang Q, et al. The mutational constraint spectrum quantified from variation in 141,456 humans. Nature. 2020;581(7809):434–43. Epub 20200527. doi: 10.1038/s41586-020-2308-7. PubMed PMID: 32461654; PubMed Central PMCID: PMCPMC7334197.

21. Cornish AJ, Gruber AJ, Kinnersley B, Chubb D, Frangou A, Caravagna G, et al. The genomic landscape of 2,023 colorectal cancers. Nature. 2024;633(8028):127–36. Epub 20240807. doi: 10.1038/s41586-024-07747-9. PubMed PMID: 39112709; PubMed Central PMCID: PMCPMC11374690.

22. Georgeson P, Harrison TA, Pope BJ, Zaidi SH, Qu C, Steinfelder RS, et al. Identifying colorectal cancer caused by biallelic MUTYH pathogenic variants using tumor mutational signatures. Nat Commun. 2022;13(1):3254. Epub 20220606. doi: 10.1038/s41467-022-30916-1. PubMed PMID: 35668106; PubMed Central PMCID: PMCPMC9170691.

23. Fang H, Zhu X, Yang H, Oh J, Barbour JA, Wong JWH. Deficiency of replication-independent DNA mismatch repair drives a 5-methylcytosine deamination mutational signature in cancer. Sci Adv. 2021;7(45):eabg4398. Epub 20211103. doi: 10.1126/sciadv.abg4398. PubMed PMID: 34730999; PubMed Central PMCID: PMCPMC8565909.

24. Degasperi A, Zou X, Amarante TD, Martinez-Martinez A, Koh GCC, Dias JML, et al. Substitution mutational signatures in whole-genome-sequenced cancers in the UK population. Science. 2022;376(6591). Epub 20220422. doi: 10.1126/science.abl9283. PubMed PMID: 35949260; PubMed Central PMCID: PMCPMC7613262.

25. Mendelaar PAJ, Smid M, van Riet J, Angus L, Labots M, Steeghs N, et al. Whole genome sequencing of metastatic colorectal cancer reveals prior treatment effects and specific metastasis features. Nat Commun. 2021;12(1):574. Epub 20210125. doi: 10.1038/s41467-020-20887-6. PubMed PMID: 33495476; PubMed Central PMCID: PMCPMC7835235.

26. Palles C, Cazier JB, Howarth KM, Domingo E, Jones AM, Broderick P, et al. Germline mutations affecting the proofreading domains of POLE and POLD1 predispose to colorectal adenomas and carcinomas. Nat Genet. 2013;45(2):136–44. Epub 20121223. doi: 10.1038/ng.2503. PubMed PMID: 23263490; PubMed Central PMCID: PMCPMC3785128.

27. Sasani TA, Ashbrook DG, Beichman AC, Lu L, Palmer AA, Williams RW, et al. A natural mutator allele shapes mutation spectrum variation in mice. Nature. 2022;605(7910):497–502. Epub 20220511. doi: 10.1038/s41586-022-04701-5. PubMed PMID: 35545679; PubMed Central PMCID: PMCPMC9272728.

28. Gray R, Wheatley K. How to avoid bias when comparing bone marrow transplantation with chemotherapy. Bone Marrow Transplant. 1991;7 Suppl 3:9–12. PubMed PMID: 1855097.

29. Katan MB. Apolipoprotein E isoforms, serum cholesterol, and cancer. Lancet. 1986;1(8479):507–8. doi: 10.1016/s0140-6736(86)92972-7. PubMed PMID: 2869248.

30. Burgess S, Davey Smith G, Davies NM, Dudbridge F, Gill D, Glymour MM, et al. Guidelines for performing Mendelian randomization investigations: update for summer 2023. Wellcome Open Res. 2019;4:186. Epub 20230804. doi: 10.12688/wellcomeopenres.15555.3. PubMed PMID: 32760811; PubMed Central PMCID: PMCPMC7384151.

31. Ashton KA, Proietto A, Otton G, Symonds I, Scott RJ. Genetic variants in MUTYH are not associated with endometrial cancer risk. Hered Cancer Clin Pract. 2009;7(1):3. Epub 20090126. doi: 10.1186/1897-4287-7-3. PubMed PMID: 19338676; PubMed Central PMCID: PMCPMC2653716.

32. Kondrashova O, Shamsani J, O’Mara TA, Newell F, McCart Reed AE, Lakhani SR, et al. Tumor Signature Analysis Implicates Hereditary Cancer Genes in Endometrial Cancer Development. Cancers (Basel). 2021;13(8). Epub 20210407. doi: 10.3390/cancers13081762. PubMed PMID: 33917078; PubMed Central PMCID: PMCPMC8067736.

33. Lintas C, Canalis B, Azzarà A, Sabarese G, Perrone G, Gurrieri F. Exploring the Role of the MUTYH Gene in Breast, Ovarian and Endometrial Cancer. Genes (Basel). 2024;15(5). Epub 20240426. doi: 10.3390/genes15050554. PubMed PMID: 38790183; PubMed Central PMCID: PMCPMC11120896.

34. Thet M, Plazzer JP, Capella G, Latchford A, Nadeau EAW, Greenblatt MS, et al. Phenotype Correlations With Pathogenic DNA Variants in the MUTYH Gene: A Review of Over 2000 Cases. Hum Mutat. 2024;2024:8520275. Epub 20240927. doi: 10.1155/2024/8520275. PubMed PMID: 40225933; PubMed Central PMCID : PMCPMC11918913.

35. Thompson AB, Sutcliffe EG, Arvai K, Roberts ME, Susswein LR, Marshall ML, et al. Monoallelic MUTYH pathogenic variants ascertained via multi-gene hereditary cancer panels are not associated with colorectal, endometrial, or breast cancer. Familial cancer. 2022;21(4):415–22. Epub 20220104. doi: 10.1007/s10689-021-00285-7. PubMed PMID: 34981295.

36. Villy MC, Masliah-Planchon J, Buecher B, Beaulaton C, Vincent-Salomon A, Stoppa-Lyonnet D, et al. Endometrial cancer may be part of the MUTYH-associated polyposis cancer spectrum. Eur J Med Genet. 2022;65(1):104385. Epub 20211111. doi: 10.1016/j.ejmg.2021.104385. PubMed PMID: 34775073.

37. Win AK, Cleary SP, Dowty JG, Baron JA, Young JP, Buchanan DD, et al. Cancer risks for monoallelic MUTYH mutation carriers with a family history of colorectal cancer. International journal of cancer. 2011;129(9):2256–62. Epub 20110408. doi: 10.1002/ijc.25870. PubMed PMID: 21171015; PubMed Central PMCID: PMCPMC3291738.

38. Zhang Y, Newcomb PA, Egan KM, Titus-Ernstoff L, Chanock S, Welch R, et al. Genetic polymorphisms in base-excision repair pathway genes and risk of breast cancer. Cancer Epidemiol Biomarkers Prev. 2006;15(2):353–8. doi: 10.1158/1055-9965.Epi-05-0653. PubMed PMID: 16492928.

39. Kaplanis J, Ide B, Sanghvi R, Neville M, Danecek P, Coorens T, et al. Genetic and chemotherapeutic influences on germline hypermutation. Nature. 2022;605(7910):503–8. Epub 20220511. doi: 10.1038/s41586-022-04712-2. PubMed PMID: 35545669; PubMed Central PMCID: PMCPMC9117138.

40. Weren RD, Ligtenberg MJ, Kets CM, de Voer RM, Verwiel ET, Spruijt L, et al. A germline homozygous mutation in the base-excision repair gene NTHL1 causes adenomatous polyposis and colorectal cancer. Nat Genet. 2015;47(6):668–71. Epub 20150504. doi: 10.1038/ng.3287. PubMed PMID: 25938944.

41. Weren RD, Ligtenberg MJ, Geurts van Kessel A, De Voer RM, Hoogerbrugge N, Kuiper RP. NTHL1 and MUTYH polyposis syndromes: two sides of the same coin? J Pathol. 2018;244(2):135–42. Epub 20171214. doi: 10.1002/path.5002. PubMed PMID: 29105096.

42. Grolleman JE, de Voer RM, Elsayed FA, Nielsen M, Weren RDA, Palles C, et al. Mutational Signature Analysis Reveals NTHL1 Deficiency to Cause a Multi-tumor Phenotype. Cancer Cell. 2019;35(2):256–66.e5. doi: 10.1016/j.ccell.2018.12.011. PubMed PMID: 30753826.

43. Palles C, West HD, Chew E, Galavotti S, Flensburg C, Grolleman JE, et al. Germline MBD4 deficiency causes a multi-tumor predisposition syndrome. Am J Hum Genet. 2022;109(5):953–60. Epub 20220422. doi: 10.1016/j.ajhg.2022.03.018. PubMed PMID: 35460607; PubMed Central PMCID: PMCPMC9118112.

44. Villy MC, Le Ven A, Le Mentec M, Masliah-Planchon J, Houy A, Bièche I, et al. Familial uveal melanoma and other tumors in 25 families with monoallelic germline MBD4 variants. J Natl Cancer Inst. 2024;116(4):580–7. doi: 10.1093/jnci/djad248. PubMed PMID: 38060262.

45. Sun JX, He Y, Sanford E, Montesion M, Frampton GM, Vignot S, et al. A computational approach to distinguish somatic vs. germline origin of genomic alterations from deep sequencing of cancer specimens without a matched normal. PLoS Comput Biol. 2018;14(2):e1005965. Epub 20180207. doi: 10.1371/journal.pcbi.1005965. PubMed PMID: 29415044; PubMed Central PMCID: PMCPMC5832436.

46. Zehir A, Benayed R, Shah RH, Syed A, Middha S, Kim HR, et al. Mutational landscape of metastatic cancer revealed from prospective clinical sequencing of 10,000 patients. Nat Med. 2017;23(6):703–13. Epub 20170508. doi: 10.1038/nm.4333. PubMed PMID: 28481359; PubMed Central PMCID: PMCPMC5461196.

47. Kinzler KW, Vogelstein B. Cancer-susceptibility genes. Gatekeepers and caretakers. Nature. 1997;386(6627):761, 3. doi: 10.1038/386761a0. PubMed PMID: 9126728.

48. Tomlinson I, Bodmer W. Selection, the mutation rate and cancer: ensuring that the tail does not wag the dog. Nat Med. 1999;5(1):11–2. doi: 10.1038/4687. PubMed PMID: 9883827.

49. Moore L, Cagan A, Coorens THH, Neville MDC, Sanghvi R, Sanders MA, et al. The mutational landscape of human somatic and germline cells. Nature. 2021;597(7876):381–6. Epub 20210825. doi: 10.1038/s41586-021-03822-7. PubMed PMID: 34433962.

50. Kidd JM, Newman TL, Tuzun E, Kaul R, Eichler EE. Population stratification of a common APOBEC gene deletion polymorphism. PLoS Genet. 2007;3(4):e63. doi: 10.1371/journal.pgen.0030063. PubMed PMID: 17447845; PubMed Central PMCID: PMCPMC1853121.

51. Xuan D, Li G, Cai Q, Deming-Halverson S, Shrubsole MJ, Shu XO, et al. APOBEC3 deletion polymorphism is associated with breast cancer risk among women of European ancestry. Carcinogenesis. 2013;34(10):2240–3. Epub 20130528. doi: 10.1093/carcin/bgt185. PubMed PMID: 23715497; PubMed Central PMCID: PMCPMC3786378.

52. Nik-Zainal S, Wedge DC, Alexandrov LB, Petljak M, Butler AP, Bolli N, et al. Association of a germline copy number polymorphism of APOBEC3A and APOBEC3B with burden of putative APOBEC-dependent mutations in breast cancer. Nat Genet. 2014;46(5):487–91. Epub 20140413. doi: 10.1038/ng.2955. PubMed PMID: 24728294; PubMed Central PMCID: PMCPMC4137149.

53. Göhler S, Da Silva Filho MI, Johansson R, Enquist-Olsson K, Henriksson R, Hemminki K, et al. Impact of functional germline variants and a deletion polymorphism in APOBEC3A and APOBEC3B on breast cancer risk and survival in a Swedish study population. J Cancer Res Clin Oncol. 2016;142(1):273–6. Epub 20150831. doi: 10.1007/s00432-015-2038-7. PubMed PMID: 26320772.

54. Middlebrooks CD, Banday AR, Matsuda K, Udquim KI, Onabajo OO, Paquin A, et al. Association of germline variants in the APOBEC3 region with cancer risk and enrichment with APOBEC-signature mutations in tumors. Nat Genet. 2016;48(11):1330–8. Epub 20160919. doi: 10.1038/ng.3670. PubMed PMID: 27643540; PubMed Central PMCID: PMCPMC6583788.

55. Gansmo LB, Romundstad P, Hveem K, Vatten L, Nik-Zainal S, Lønning PE, et al. APOBEC3A/B deletion polymorphism and cancer risk. Carcinogenesis. 2018;39(2):118–24. doi: 10.1093/carcin/bgx131. PubMed PMID: 29140415; PubMed Central PMCID: PMCPMC5862322.

56. Hashemi M, Moazeni-Roodi A, Taheri M. Association of APOBEC3 deletion with cancer risk: A meta-analysis of 26 225 cases and 37 201 controls. Asia Pac J Clin Oncol. 2019;15(6):275–87. Epub 20190129. doi: 10.1111/ajco.13107. PubMed PMID: 30693645.

57. Mouradov D, Greenfield P, Li S, In EJ, Storey C, Sakthianandeswaren A, et al. Oncomicrobial Community Profiling Identifies Clinicomolecular and Prognostic Subtypes of Colorectal Cancer. Gastroenterology. 2023;165(1):104–20. Epub 20230316. doi: 10.1053/j.gastro.2023.03.205. PubMed PMID: 36933623.

58. Bjelakovic G, Nikolova D, Simonetti RG, Gluud C. Antioxidant supplements for prevention of gastrointestinal cancers: a systematic review and meta-analysis. Lancet. 2004;364(9441):1219–28. doi: 10.1016/s0140-6736(04)17138-9. PubMed PMID: 15464182.

59. Raczy C, Petrovski R, Saunders CT, Chorny I, Kruglyak S, Margulies EH, et al. Isaac: ultra-fast whole-genome secondary analysis on Illumina sequencing platforms. Bioinformatics. 2013;29(16):2041–3. Epub 20130604. doi: 10.1093/bioinformatics/btt314. PubMed PMID: 23736529.

60. Poplin R, Chang PC, Alexander D, Schwartz S, Colthurst T, Ku A, et al. A universal SNP and small-indel variant caller using deep neural networks. Nat Biotechnol. 2018;36(10):983–7. Epub 20180924. doi: 10.1038/nbt.4235. PubMed PMID: 30247488.

61. McLaren W, Gil L, Hunt SE, Riat HS, Ritchie GR, Thormann A, et al. The Ensembl Variant Effect Predictor. Genome Biol. 2016;17(1):122. Epub 20160606. doi: 10.1186/s13059-016-0974-4. PubMed PMID: 27268795; PubMed Central PMCID: PMCPMC4893825.

62. Landrum MJ, Lee JM, Benson M, Brown GR, Chao C, Chitipiralla S, et al. ClinVar: improving access to variant interpretations and supporting evidence. Nucleic Acids Res. 2018;46(D1):D1062–d7. doi: 10.1093/nar/gkx1153. PubMed PMID: 29165669; PubMed Central PMCID: PMCPMC5753237.

63. Jaganathan K, Kyriazopoulou Panagiotopoulou S, McRae JF, Darbandi SF, Knowles D, Li YI, et al. Predicting Splicing from Primary Sequence with Deep Learning. Cell. 2019;176(3):535–48.e24. Epub 20190117. doi: 10.1016/j.cell.2018.12.015. PubMed PMID: 30661751.

64. Nik-Zainal S, Alexandrov LB, Wedge DC, Van Loo P, Greenman CD, Raine K, et al. Mutational processes molding the genomes of 21 breast cancers. Cell. 2012;149(5):979–93. Epub 20120517. doi: 10.1016/j.cell.2012.04.024. PubMed PMID: 22608084; PubMed Central PMCID: PMCPMC3414841.

65. Van Loo P, Nordgard SH, Lingjærde OC, Russnes HG, Rye IH, Sun W, et al. Allele-specific copy number analysis of tumors. Proc Natl Acad Sci U S A. 2010;107(39):16910–5. Epub 20100913. doi: 10.1073/pnas.1009843107. PubMed PMID: 20837533; PubMed Central PMCID: PMCPMC2947907.

66. Salipante SJ, Scroggins SM, Hampel HL, Turner EH, Pritchard CC. Microsatellite instability detection by next generation sequencing. Clin Chem. 2014;60(9):1192–9. Epub 20140630. doi: 10.1373/clinchem.2014.223677. PubMed PMID: 24987110.

67. Rayner E, van Gool IC, Palles C, Kearsey SE, Bosse T, Tomlinson I, et al. A panoply of errors: polymerase proofreading domain mutations in cancer. Nat Rev Cancer. 2016;16(2):71–81. doi: 10.1038/nrc.2015.12. PubMed PMID: 26822575.

68. Bergstrom EN, Huang MN, Mahto U, Barnes M, Stratton MR, Rozen SG, et al. SigProfilerMatrixGenerator: a tool for visualizing and exploring patterns of small mutational events. BMC Genomics. 2019;20(1):685. Epub 20190830. doi: 10.1186/s12864-019-6041-2. PubMed PMID: 31470794; PubMed Central PMCID: PMCPMC6717374.

69. Díaz-Gay M, Vangara R, Barnes M, Wang X, Islam SMA, Vermes I, et al. Assigning mutational signatures to individual samples and individual somatic mutations with SigProfilerAssignment. Bioinformatics. 2023;39(12). doi: 10.1093/bioinformatics/btad756. PubMed PMID: 38096571; PubMed Central PMCID: PMCPMC10746860.

